# Convergent Innate Immune and Metabolic Signatures in Parkinson’s Disease and Viral Infection

**DOI:** 10.64898/2026.08.28.26361092

**Authors:** Megan Belyea, Mohsin Shafiq, Joshua Laß, Christoph Much, Zhezhang Liu, Nathalie Kruse, Kristian Händler, Varun Sreenivasan, Ellen Gelpi, Bente Siebels, Benjamin Ondruschka, Malte Spielmann, Christine Klein, Joanne Trinh, Markus Glatzel

**Author notes:** **Corresponding author information:** Markus Glatzel, MD, Institute of Neuropathology, University Medical Center Hamburg-Eppendorf Martinistrasse 52, 20246, Hamburg, Germany. equal contribution (Megan Belyea, Mohsin Shafiq, Joanne Trinh, Markus Glatzel).

## Abstract

Viral infections have long been proposed as environmental contributors to neurodegenerative diseases, including Parkinson’s disease (PD), yet the molecular mechanisms linking infection and neurodegeneration are not well defined. Neuroinflammation and disruption of central nervous system (CNS) homeostasis have emerged as potential mediators. In this study, we used severe acute respiratory syndrome coronavirus 2 (SARS-CoV-2), the causative agent of COVID-19, as a model pathogen to investigate convergent molecular pathways between viral infection and PD. Single-nucleus RNA sequencing (snRNA-seq) was performed on post-mortem striatal tissue from 14 individuals stratified into four groups: COVID-19 only (COVID-19), PD only (PD), comorbid PD with COVID-19 (PD/COVID-19), and controls (Control). The PD/COVID-19 group exhibited an expanded astrocytic population and a pronounced interferon-associated molecular signature characterized by increased expression of canonical interferon-stimulated genes, including *IFI44L* (average log2FC= 3.9; adjusted p=2.3 x 10^−373^)*, IFI44* (average log2FC=2.9; adjusted p=8.0 x 10^−266^)*, ISG15* (average log2FC=3.1; adjusted p=1.2 x 10^−197^), and *RSAD2* (average log2FC= 3.5; adjusted p=8.6 x 10^−111^). Pathway analyses demonstrated activation of innate immune and antiviral signaling pathways, particularly within microglia and astrocytes, including interferon signaling, pattern-recognition receptor pathways, and complement-associated responses. In parallel, genes involved in lipid metabolism, cholesterol homeostasis, synaptic maintenance, and neuronal signaling were reduced across disease groups. Proteomic analyses independently confirmed enrichment of antiviral and interferon-associated pathways and identified convergent suppression of sterol, cholesterol, and lipid metabolic processes. Our findings identify a convergent molecular signature linking PD and COVID-19, pronounced in comorbid individuals and characterized by interferon-driven innate immune activation, glial inflammatory responses, and dysregulation of lipid metabolic homeostasis. Collectively, the data support a model in which severe viral infection amplifies biological pathways already implicated in PD pathogenesis.

## Introduction

Parkinson’s disease (PD) is a progressive neurodegenerative disorder characterized by dopaminergic neuronal loss, α-synuclein aggregation, motor dysfunction, and a broad spectrum of non-motor symptoms. Mechanisms driving disease manifestation are not fully understood [8]. In addition to genetic susceptibility, environmental factors are increasingly recognized as important disease drivers [7]. Among these, infectious diseases, particularly viral infections, have long been proposed as potential modulators of neurodegenerative processes [10, 16]. Epidemiological observations following the 1918 influenza pandemic reported increased rates of post-encephalitic parkinsonism and suggested a potential association between severe viral infection and later neurodegenerative disease [44]. Cohort analyses have reinforced this pattern, indicating that individuals born during this period exhibited a two-to-three-fold higher lifetime risk of idiopathic PD [36]. More recently, large population-based studies have demonstrated associations between viral infections and increased risks of neurodegenerative diseases [30, 33, 46]. Similarly, recent meta-analyses have reported associations between several viral pathogens and increased risks of both Alzheimer’s disease and PD [33], suggesting that infection-associated neurodegeneration may represent a broader biological phenomenon rather than a pathogen-specific effect.

Several mechanisms have been proposed to explain these associations. Experimental and translational studies suggest that viral infections can induce persistent neuroinflammatory responses, disrupt protein homeostasis, alter cellular metabolism, and impair neuronal function [27, 38]. These processes are highly relevant to PD pathogenesis, where chronic activation of innate immune pathways, glial dysfunction, oxidative stress, and impaired proteostasis are increasingly recognized as major contributors to neuronal vulnerability [12]. Rather than acting through direct viral neurotoxicity, infections may alter neurodegenerative disease by inducing neuroinflammation, microglial activation, and altering proteostasis. Support for this concept comes from experimental studies with influenza strains such H5N1 [22].

The emergence of Coronavirus Disease 2019 (COVID-19), caused by severe acute respiratory syndrome coronavirus 2 (SARS-CoV-2), provides a unique opportunity to investigate these mechanisms. Beyond its respiratory manifestations, COVID-19 is associated with a wide range of neurological complications, including cognitive impairment, anosmia, cerebrovascular abnormalities, white matter damage and neuropsychiatric symptoms [9, 47]. Neuropathological studies have demonstrated widespread neuroimmune activation characterized by microgliosis, astrogliosis, inflammatory signaling, vascular alterations, and disturbances of neuronal homeostasis [19, 49, 61]. These observations have raised concerns regarding the potential long-term neurological consequences of severe SARS-CoV-2 infection.

Particular attention has focused on individuals with pre-existing neurodegenerative disorders. Patients with PD experience increased morbidity, hospitalization rates, and mortality following COVID-19 and frequently report worsening of motor and non-motor symptoms during and after infection [3, 48, 54, 55]. While these observations suggest that PD patients may be particularly vulnerable to systemic viral insults, the molecular basis of this vulnerability remains unclear. Importantly, whether SARS-CoV-2 activates pathways that converge with established mechanisms of PD pathogenesis has not been systematically investigated at single-cell resolution in human brain tissue.

Here, we used SARS-CoV-2 infection as a model of severe viral disease to investigate molecular convergence between viral infection and PD. We performed single-nucleus RNA sequencing of post-mortem striatal tissue from individuals with PD, PD/COVID-19, COVID-19, and controls. Transcriptomic findings were complemented by proteomic analyses of the same individuals. By identifying shared and distinct molecular signatures across these conditions, we identified interferon-driven innate immune activation, glial inflammatory responses, and dysregulation of lipid metabolic homeostasis as pronounced alterations in comorbid PD/COVID-19 and thus provide novel insight into how viral infections may modulate neurodegenerative processes and contribute to disease vulnerability.

## Methods

### Description of patients and controls

Patients who had died with a recent history of COVID-19 alone, but also patients with a recent history of COVID-19 and a concomitant diagnosis of PD and patients without COVID-19 and PD were autopsied at the Institute of Legal Medicine at the University Medical Center of Hamburg-Eppendorf. Additionally, patients with PD diagnosed at the Division of Neuropathology and Neurochemistry, Department of Neurology, Medical University of Vienna, were included in the study. Ethical approval for the use of postmortem brain tissue for research studies was obtained from both Institutions (MedUniWien EK1454/2018, 2020-10353-BO-ff). Prior to fixation in buffered 4% formaldehyde, cryosamples from the substantia nigra and the striatum were taken and processed for single-nuclei sequencing. Brains were examined macroscopically and underwent routine neuropathological workup to categorize PD and the typical SARS-CoV-2 infection changes as published [5, 37]. Participant demographics and pathology are summarized in Table 1. Individuals (n=16) were placed into four different cohorts (n=4 in each) based on disease status: individuals with COVID-19 but not PD (COVID-19), individuals with PD but not COVID-19 (PD), individuals with both PD and COVID-19 (PD/COVID-19), and individuals without SARS-CoV-2 infection and PD were the control group (Control).

**Table 1.** Clinical and neuropathological characteristics of the study cohort.

| ID | PD symptoms /<br>pathology consensus | Sex | Age<br>(Range) | COVID-19<br>duration<br>(days) | PMI<br>(days) | Cause of<br>death | Comorbidities | Brain details:<br>weight (g) / edema/<br>atrophy | Macroscopic findings |
| --- | --- | --- | --- | --- | --- | --- | --- | --- | --- |
| COVID-19-1 | No | m | ≥70 | 8 | 3 | PNA | Obesity, COPD, CI, severe AS | 1515 / Mild / None | None |
| COVID-19-2 | No | f | ≥70 | 5 | 4 | Subdural<br>hemorrhage | PNA, CI, I, mild AS | 1235 / Mild / None | Acute subdural<br>hemorrhage |
| COVID-19-3 | No | f | ≥70 | 17 | 5 | PNA | CI, RI, mild AS | 1280 / None / Mild | Old cerebellar infarction |
| COVID-19-4 | No | f | ≥70 | 5 | 3 | PNA | CI, RI, dementia, severe AS | 1380 / None / Mild | None |
| PD-1 | Yes / DLB/PD (neocortical) | m | ≤69 | n/a | 3 | Paralytic<br>ileus, PNA | PE, AS | 1350 / None / None | Marked symmetric<br>pallor SN and LC |
| PD-2 | Yes / DLB/PD (neocortical) | m | ≥70 | n/a | 2 | MI | HT, HC, HI, CI, RI, AS | 1297 / Mild / Mild | Marked symmetric<br>pallor SN and LC |
| PD-3 | Yes / DLB/PD (neocortical) | m | ≥70 | n/a | 2 | PE | NA | 1283 / None / Mild | Marked pallor SN and LC |
| PD-4 | Yes / DLB/PD (neocortical) | m | ≥70 | n/a | NA | PNA | NA | NA / Mild / None | Moderate symmetric<br>pallor SN and LC |
| PD/COVID-19-1 | Yes / DLB/PD (brainstem<br>predominant) | m | ≤69 | 3 | 4 | MI | CI, COPD, mild AS | 1650 / Mild / None | None |
| PD/COVID-19-2 | Yes / DLB/PD (brainstem<br>predominant) | m | ≥70 | 24 | 4 | PNA | CI, RI, mild AS | 1250 / None / None | None |
| PD/COVID-19-3 | Yes / DLB/PD (brainstem<br>predominant) | m | ≥70 | 14 | 2 | PNA | COPD, severe AS | 1355 / Mild / None | None |
| PD/COVID-19-4 | Yes / DLB/PD (neocortical) | m | ≤69 | 7 | 2 | PNA | CI, COPD, mild AS | 1400 / Mild / None | None |
| Control-1 | No | f | ≥70 | n/a | 3 | MI | CI, moderate AS | 1210 / None / None | None |
| Control-2 | No | m | ≤69 | n/a | 4 | PNA | Liver Tx, hep B, moderate AS | 1215 / Mild / None | None |
| Control-3 | No | m | ≥70 | n/a | 2 | PE | CI, mild AS | 1235 / None / None | Meningioma |
| Control-4 | No | f | ≥70 | n/a | 4 | Suffocation | COPD, CI, DM | 1310 / Mild / Mild | None |

*Legend: m: male, f: female; n/a: not available; cause of death: PE: pulmonary artery embolisms, MI: myocardial infarction; PMI: postmortem interval; co-morbidities: CI: cardiac insufficiency, COPD: chronic obstructive pulmonary disease, DM: diabetes mellitus, RI: renal insufficiency, Tx: Transplantation, PNA: Pneumonia; AS: arteriosclerosis; HT: Hypertension; HC: hypercholesterolemia, hep B: hepatitis B*

### Morphological analyses

Patients included in this study underwent neuropathological examination to confirm the diagnosis of PD and exclude other neuropathological alterations. Briefly, formalin-fixed paraffin-embedded tissue (FFPE) samples from neocortical regions, the amygdala, the brainstem, the limbic system and the cerebellum were processed and stained according to current neuropathological neurodegeneration grading schemes [5, 39].

Representative histological images from the striatum, the region where single-nuclei isolation was performed, are shown. For this, haematoxylin and eosin (HE) staining were performed using standard laboratory procedures (Figure 1, Supplementary Figure 1). Immunohistochemical staining was achieved using a Ventana Benchmark XT Autostainer (Ventana, Tucson, AZ, USA), in accordance with the manufacturer’s recommendations, using antibodies against human glial fibrillary acidic protein (GFAP; clone 6F2; Dako, Glostrup, Denmark; dilution 1:200), HLA-DR (mouse anti-HLA-DP, DQ, DR antibody, clone CR3/43; Dako; 1:200) and a-synuclein (Zytomed 519-2684; 1:1000). Slides were electronically scanned at high magnification (×40) as high-resolution images (1900 × 1200 pixels) with a NanoZoomer 2.0-HT (Hamamatsu Photonics, Hamamatsu, Japan).

**Figure 1.**
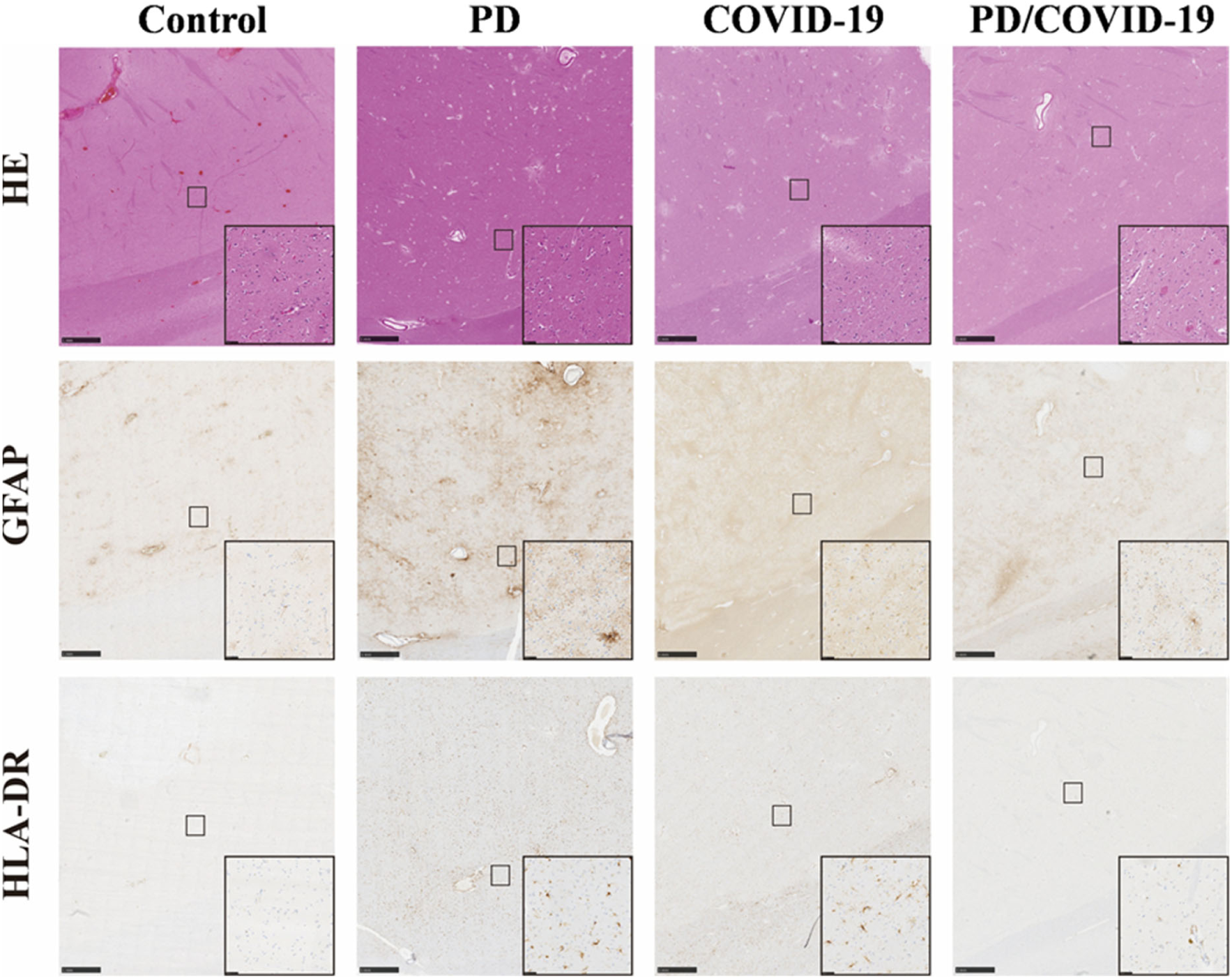
Histological findings in the striatum of patients with PD/COVID-19, COVID-19, PD, and controls. Representative images of haematoxylin and eosin (HE) staining are shown in the first row. GFAP immunohistochemistry (second row) demonstrated reactive astrogliosis in PD/COVID-19, COVID-19, and PD cases, with less prominent activation in controls. HLA-DR immunohistochemistry (third row) showed microglial activation in PD/COVID-19 and COVID-19 cases, less prominent activation in PD, and no activation in controls. Scale bars represent 1 mm in the main images and 50 μm in the insets

### Single-nuclei isolation

Single-nuclei RNA sequencing by 10X Genomics was performed on the striatum of 16 individuals categorized as COVID-19, PD, PD/COVID-19 and Control (n = 4 for each group). Striatal tissue was extracted from post-mortem brains and directly frozen in liquid nitrogen. Single-nuclei experiments were performed in accordance with a newly developed and optimized 10X Genomics protocol (as outlined by Much et al. 2025 [40]) for single-nuclei gene expression and Chromium fixed RNA profiling [1], requiring an initial amount of tissue of 30 mg. In brief, each frozen sample was homogenized and incubated for 10-minutes in lysis buffer solution prior to being transferred into a nuclei isolation column for centrifugation, washing and debris removal, and nuclei collection. Fluorescence-Activated Cell Sorting (FACs) was used to filter isolated nuclei, and only single DAPI-positive cells were used in our study. The library was prepared using the unique human whole transcriptome atlas probe barcode prior to sequencing on the NextSeq 2000 system from Illumina.

### Single-nucleus RNA-seq data processing and analysis

Cell identification and sample demultiplexing were performed using Cell Ranger (version 7.0.0, 10X Genomics). Quality control, normalization, and clustering were performed using the Seurat (v5.4) package in RStudio. Genes detected in fewer than three nuclei were excluded. Nuclei were filtered to retain those with at least 800 total UNI counts and at least 800 detected genes, and nuclei with greater than 5% mitochondrial transcript fraction were filtered out. Doublets and multiplets were identified per sample using Scrublet (expected doublet rate 0.02), and nuclei were filtered out using the 30-PC analysis branch with a hybrid floor threshold (equal to the greater of the sample-specific Scrublet auto-threshold and 0.15).

Expression values were log-normalized, and 4,000 highly variable genes were selected. All analyses were performed using the RNA assay (RNANormalize). Samples were merged into a single Seurat object, scaled, and subjected to principal component analysis (PCA; 35 PCs). A shared nearest neighbor (SNN) graph was constructed using PCA dimensions 1–10, and clustering was performed using the Louvain algorithm at resolution 0.05. UMAP embeddings were generated using PCA reductions (uwot; cosine distance; n.neighbors=30; min.dist=0.3; seed=42). For visualization, an additional UMAP was generated from Harmony-transformed PCA embeddings (15 dimensions; nclust=12) to facilitate integrated cross-sample comparison, while PCA-based embeddings were used for downstream analyses. Initial cluster marker genes were identified using Seurat’s differential expression testing (Wilcoxon rank-sum test).

Clusters were annotated based on the top differentially expressed genes (typically top 10, expanded to top 40 where needed), prioritizing genes with high expression prevalence (pct.1) and enrichment relative to other clusters (log2 fold-change). Cell identities were assigned through comparison with curated reference databases (PanglaoDB and CellMarker 2.0) and manual literature curation.

### Differential expression analysis of single-nuclei RNA sequencing data

Differential expression analyses were performed on the RNA assay in Seurat. For analysis, the integrated Seurat object containing all cells was stratified into four condition groups: COVID-19, PD, PD/COVID-19 and Control. Differential expression testing was performed within each annotated cell type using pairwise comparisons against the control group (Control). Specifically, genes were identified for each condition (COVID-19, PD, and PD/COVID-19) relative to control (Control) (Supplementary Figure 2).

In addition to three pairwise comparisons, we included analyses to assess genes shared across COVID-19, PD, and PD/COVID-19 groups to determine whether the combined PD/COVID-19 condition was associated with increased magnitude of differential expression. Genes were classified as showing amplified comorbidity-associated effects if they changed in the same direction across all groups, exhibited a strong effect in PD/COVID-19 (|log₂FC| > 3), and showed a meaningful effect in at least one individual condition (PD or COVID-19; |log₂FC| > 1).

Differential expression testing was performed using Seurat’s FindMarkers function (Wilcoxon rank-sum test). Genes were included for testing if detected in at least 5% of cells in either comparison group (min.pct=0.05), with no log2FC pre-filter applied. Raw p-values were corrected for multiple testing using the Benjamini-Hochberg procedure; genes with an adjusted p-value<0.05 were considered significant. Log-normalized expression values from RNA assay were used for all differential expression analyses to ensure consistent normalization across comparisons.

For each cell type, the top 10 maximum up- and down-regulated genes were selected based on |adjusted p-value| (<0.05) and a stringent fold-change threshold of |log₂FC|>3, corresponding to a minimum 8-fold difference in expression between groups. This threshold was applied to highlight genes showing the most pronounced changes, while a complete list of differentially expressed genes across all thresholds is provided in Supplementary Tables 1-3.

### Pathway enrichment analysis of single-nuclei RNA data

Exploratory pathway enrichment analysis was performed using pre-ranked gene set enrichment analysis (GSEA) on differential expression results derived from the RNA assay of the annotated Seurat object. Differential expression was computed across all cells and within each annotated cell type for the pairwise baseline contrasts, as previously described, using the Wilcoxon rank-sum test in Seurat. Genes were ranked using a signed score reflecting both the direction (log2 fold-change) and statistical significance of differential expression. The full set of tested genes was retained for pathway analysis. KEGG gene sets were obtained from MSigDB via the msigdbr R package (KEGG), and enrichment was performed using the fgsea package in human gene-symbol space. Gene sets containing 10–500 genes were tested. Positive normalized enrichment scores indicated enrichment among genes upregulated in the disease group relative to control, whereas negative scores indicated enrichment among relatively downregulated genes.

### Tryptic Digestion for Mass spectrometry-based bottom-up proteomics

Samples were dissolved in 100 mM triethyl ammonium bicarbonate (TEAB) and 1% w/v sodium deoxycholate (SDC) buffer, boiled at 95 °C for 5 min and sonicated with a probe sonicator. The protein concentration of denatured proteins was determined by the Pierce bicinchoninic acid assay (BCA) Protein assay kit (Thermo Fisher) and samples were diluted to 10 µg of protein. The digestion was performed in a 96-well LoBind plate (Eppendorf, Hamburg, Germany) semi-automated on an Andrew+ Pipetting Robot (Waters, Milford, USA). Disulfide bonds were reduced in 10 mM dithiothreitol for 30 min at 56 °C while shaking at 800 rpm and alkylated in presence of 20 mM iodoacetamide for 30 min at 37 °C. Carboxylate modified magnetic E3 and E7 speed beads (Cytvia Sera-Mag™, Marlborough, USA) at 1:1 ratio in LC-MS grade water were added in a 10:1 (beads/protein) ratio adapted from the SP3-protocol workflow [21]. Protein binding was performed at 50% ACN during shaking at 600 rpm for 18 mins. Magnetic beads were washed two times with 80% Ethanol (EtOH) and 100% ACN. Digestion with trypsin in 100 mM AmBiCa was performed (sequencing grade, Promega) at 1:100 (enzyme:protein) ratio at 37 °C overnight while shaking at 500 rpm. Trifluoroacetic acid (TFA) was added to a final concentration of 1% to inactivate trypsin and shaken at 500 rpm for 5 min. The supernatant containing tryptic peptides was transferred into a new 96-well LoBind plate, ready for subsequent LC-MS/MS analysis.

### LC-MS/MS measurement

Chromatographic separation of tryptic peptides was achieved with a two-buffer system (buffer A: 0.1% formic acid (FA) in H2O, buffer B: 0.1% FA in 80% ACN) on a UHPLC (VanquishTM neo UHPLC system, Thermo Fisher) at a flow rate of 0.4 µL/min. Attached to the UHPLC was a peptide trap (300 µm x 5 mm, C18, PepMap™ Neo Trap Cartridge, Thermo Fisher) for online desalting and purification, followed by a 25 cm C18 reversed-phase column (120 Å, 1,7 µm, 75 µm x 250 mm, Aurora Ultimate, IonOptics). Peptides were separated using a 70 min method with linearly increasing ACN concentration from 0 to 4 % buffer B in 1 min and increased to 43 % in 60 min.

MS/MS measurements were performed on a quadrupole-orbitrap hybrid mass spectrometer (Exploris 480, Thermo Fisher Scientific). Eluting peptides were ionized using a nano-electrospray ionization source (nano-ESI) with a spray voltage of 1,800 and analyzed in data independent acquisition (DIA) mode. For each MS1 scan, ion accumulation time was set to automatic, with AGC target of 300%. The scan was set to m/z 400-800 with a resolution of 120,000 at m/z 200. Within a precursor mass range of m/z 400-800 fragmentation in DIA-mode with m/z 12 isolation windows and m/z 1 window overlaps was performed (total of 33 scan events). Fragmentation was performed at normalized collision energy of 30% using higher energy collisional dissociation (HCD). Orbitrap resolution was set to 60 000.

### LC-MS/MS data processing

LC-MS/MS data were searched with the CHIMERYS DIA algorithm integrated into the Proteome Discoverer software (v3.1.0.638, Thermo Fisher Scientific) against a reviewed human Swissprot database using Inferys 3.0 fragmentation as prediction model. Carbamidomethylation was set as a fixed modification for cysteine residues. The oxidation of methionine was allowed as variable modification. A maximum number of one missing tryptic cleavage was set. Peptides between 7 and 30 amino acids were considered. A strict cutoff (FDR < 0.01) was set for peptide and protein identification. Quantification was performed by CHIMERYS based on fragment ions. The mass spectrometry proteomics data have been deposited to the ProteomeXchange Consortium via the PRIDE (partner repository with the dataset identifier PXD080847).

### Statistical and bioinformatics analysis of proteomics data

Obtained protein abundances were log2-transformed and normalized to the column median in Perseus [56]. Normalized data were imported into R and analyzed using base R and Bioconductor/CRAN packages. The dataset contained four experimental groups: Control, PD, COVID, and PD/COVID-19. For group-wise detection analyses, proteins were retained when they contained no more than two missing values within a given group. Remaining missing values were imputed using the corresponding row median within each group. Protein overlap across groups was assessed using group-specific detected protein lists. Total detected proteins, uniquely detected proteins, and shared protein subsets were summarized using Venn membership tables and visualized using bar plots.

For unsupervised analyses, proteins with remaining missing values were excluded. Principal component analysis was performed using the prcomp function in base R after transposition of the abundance matrix so that samples represented observations. Sample–sample Pearson’s correlation matrices were calculated using pairwise complete observations and visualized as clustered heatmaps. Heatmaps of protein abundance patterns were generated using row-wise scaling. For global group effects, one-way ANOVA was applied protein-wise across the four groups.

Pairwise differential abundance analyses were performed for PD versus Control, COVID versus Control, and PD/COVID-19 versus Control comparisons. For each protein, log2 fold-change was calculated as the difference between group means, and statistical significance was assessed using two-sided Student’s t-tests. Proteins were considered differentially abundant when |Log2FC|> 0.584, corresponding to a >1.5-fold change, and the nominal p-value<0.05. P-values were adjusted for multiple testing using the Benjamini–Hochberg method and are reported in the supplementary data. Gene set enrichment analysis was performed using clusterProfiler with GO gene sets obtained from msigdbr. Proteins were ranked using a signed metric calculated as log2 fold-change multiplied by −log10(p-value). Analyses were performed for GO Biological Process gene sets using Homo sapiens annotations, with minGSSize=3, maxGSSize=500, p-valueCutoff=0.05, and Benjamini–Hochberg correction. Enriched terms were visualized using dot plots. Significantly enriched pathways were defined as those with adjusted p<0.05 and absolute normalized enrichment score > 1.5. To identify disease-associated inflammatory and infection-related signatures, differentially upregulated proteins were combined with proteins uniquely detected in each disease group, while downregulated proteins were analyzed separately. GO Biological Process overrepresentation analysis was then performed using compareCluster for the PD, COVID, and PD/COVID-19 groups. Infection- and immune-related GO terms were further extracted using keyword matching for terms related to viral processes, interferon signaling, innate immunity, antimicrobial responses, and host defense.

To assess potential comorbidity-associated amplification, proteins were evaluated across PD, COVID, and PD/COVID-19 comparisons relative to controls. Proteins were classified as amplified when they changed in the same direction in all three disease comparisons and showed a greater absolute log2 fold-change in the PD/COVID-19 group than in either PD or COVID alone. Proteins that met these criteria and showed p<0.05 in all three comparisons were retained as amplified comorbidity-associated candidates. Figures were generated using ggplot2, pheatmap, ComplexHeatmap, ggcorrplot, enrichplot, and related R packages.

### Literature Review

A structured literature search was performed in PubMed to evaluate the availability of single-cell and single-nucleus RNA sequencing studies examining PD in the context of SARS-CoV-2 infection in human post-mortem brain tissue. Studies related to PD and COVID-19 were identified using the following query: (Parkinson*[Title/Abstract]) AND (COVID-19[Title/Abstract] OR SARS-CoV-2[Title/Abstract]) AND (brain[Title/Abstract] OR CNS[Title/Abstract] OR “central nervous system”[Title/Abstract] OR postmortem[Title/Abstract] OR “post-mortem”[Title/Abstract] OR autopsy[Title/Abstract]) NOT (review[Publication Type] OR systematic review[Publication Type] OR meta-analysis[Publication Type] OR scoping review[Publication Type] OR editorial[Publication Type] OR comment[Publication Type] OR letter[Publication Type]). All retrieved records were manually screened for relevance. Studies were included if they: 1) investigated Parkinson’s disease in the context of COVID-19 or SARS-CoV-2, 2) involved human subjects or human-derived systems, 3) reported primary experimental or clinical findings. Studies were excluded if they were non-primary literature (e.g., reviews, perspectives, or computational-only studies), conducted exclusively in non-human models without human relevance, did not directly assess the interaction between PD and COVID-19, or were retracted and 4) employed single-cell or single-nucleus RNA sequencing in brain tissue or central nervous system tissue.

## Results

### Single-nuclei RNA sequencing

A total of 62,166 nuclei from 14 out of 16 patients (n=4 for COVID-19; n= 4 for PD; n=3 for PD/COVID-19; and n=3 for Control) were used for downstream analysis after filtering with Scrublet software. Two out of 16 patients were filtered out (n=1 from PD/COVID-19; and n=1 for Control) as these did not pass Scrublet QC, likely due to doublets or errors in cell sorting or capture.

### Descriptive cluster composition in each group

The highest number of nuclei were from the PD group (n= 28,844), and accounted for 46% (n=28,844/62,166) of the total nuclei. The COVID-19 group had 13,933 nuclei out of 62,166 total (22%); whereas, the control group (Control) had 13,742 nuclei out of 62,166 total (22%). The lowest number of nuclei was detected in the PD/COVID-19 group, accounting for only 9% (n= 5,647/62,166 nuclei) of the total.

Clustering identified eight distinct cell types: oligodendrocytes (n=33,234), astrocytes (n=7,788), microglia (n=4,643), oligodendrocyte precursor cells (OPCs; n=3,786), neurons (n=4,160), excitatory neurons (n=2,839), medium spiny neurons (MSNs; n=4,731), and vascular-associated cells (n=985) (Figure 2). The oligodendrocyte cluster was the largest overall (n=33,234/62,166 total nuclei; 53%). Oligodendrocytes made up 45% (n=13,064/28,844) of the PD, 64% (n=8,880/13,933) of the COVID-19, 55% (n=3,096/5,647) of the PD/COVID-19, and 60% (n=8,194/13,742) of the Control group. Astrocytes were the largest in the combined PD/COVID-19 group. The astrocyte cluster accounted for 12% (n=3,596/28,844) of the PD, 13% (n=1,854/13,933) of the COVID-19, 20% (n=1,141/5,647) of the PD/COVID-19, and 9% (n=1,197/13,742) of the Control group. Microglia were the largest cluster in the control group, making up 4% (n=1,198/28,844) of the PD, 6% (n=830/13,933) of the COVID-19, 6% (n=360/5,647) of the PD/COVID-19 and 16% (n=2,255/13,742) of the Control. OPCs accounted for 4% (n=1,274/28,844) of the PD, 7% (n=999/13,933) of the COVID-19, 10% (n=537/5,647) of the PD/COVID-19, and 7% (n=976/13,742) of the Control group. Neurons were comparable across groups: 7% (n=1,985/28,844) in the PD, 6% (n=868/13,933) in the COVID-19, 6% (n=325/5,647) in the PD/COVID-19, and 7% (n=982/13,742) in the Control group. As the PD group accounted for over 99% of the medium spiny neurons (n=4,722/4,731 cells) and excitatory neurons (n=2824/4160 cells), the comparisons for these cell types in this group were interpreted with caution. Vascular-associated cells made up 1% (n=181/28,844) of the PD, 4% (n=491/13,933) of the COVID-19 and 3% (n=181/5,647) of the PD/COVID-19 and 1% (n=132/13,742) of the Control group (Table 2).

**Figure 2.**
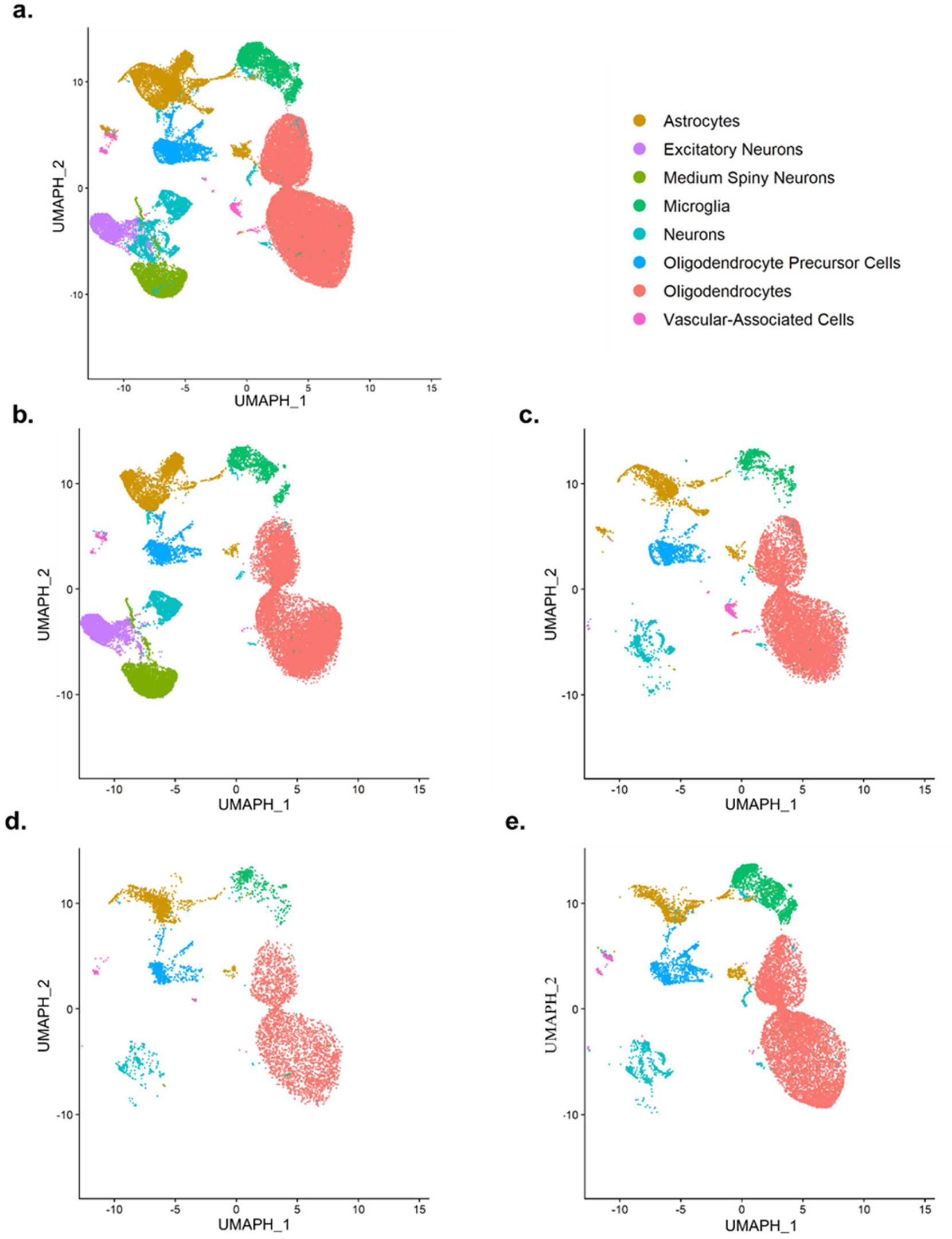
Harmony-corrected UMAP visualization of nuclei used for analysis. Nuclei (n = 62,166) from n = 14 patients were analyzed and visualized using a Harmony-corrected UMAP embedding to improve alignment of shared cell-type structure across samples. Clusters were assigned to broad biological cell types based on cluster marker gene expression from the annotated merged dataset, and colors indicate annotated cell types as shown in the legend. a. all cells from all four groups; b. PD group (PD; n=28,844); c. cells from the COVID-19 group (n=13,933); d. cells from the control group (n=13,742); e. cells from the PD/COVID-19 group (n=5,647)

**Table 2.** Distribution of nuclei across annotated cell-type clusters.

| Cell-type cluster | All nuclei<br>(N = 62,166) | Control<br>(N = 13,742) | PD<br>(N = 28,844) | COVID-19<br>(N = 13,933) | PD/COVID-19<br>(N = 5,647) |
| --- | --- | --- | --- | --- | --- |
| Astrocytes | 7,788 (12.5%) | 1,197 (8.7%) | 3,596 (12.5%) | 1,854 (13.3%) | 1,141 (20.2%) |
| Microglia | 4,643 (7.5%) | 2,255 (16.4%) | 1,198 (4.2%) | 830 (6.0%) | 360 (6.4%) |
| Neurons | 4,160 (6.7%) | 982 (7.1%) | 1,985 (6.9%) | 868 (6.2%) | 325 (5.8%) |
| Excitatory neurons | 2,839 (4.6%) | 6 (<0.1%) | 2,824 (9.8%) | 8 (0.1%) | 1 (<0.1%) |
| Medium spiny neurons | 4,731 (7.6%) | 0 (0.0%) | 4,722 (16.4%) | 3 (<0.1%) | 6 (0.1%) |
| Oligodendrocytes | 33,234 (53.5%) | 8,194 (59.6%) | 13,064 (45.3%) | 8,880 (63.7%) | 3,096 (54.8%) |
| Oligodendrocyte precursor cells | 3,786 (6.1%) | 976 (7.1%) | 1,274 (4.4%) | 999 (7.2%) | 537 (9.5%) |
| Vascular-associated cells | 985 (1.6%) | 132 (1.0%) | 181 (0.6%) | 491 (3.5%) | 181 (3.2%) |

*Data are shown as n (% of group total). Nuclei were assigned to Seurat clusters annotated using PanglaoDB and CellMarker. Percentages may not total 100% because of rounding. Abbreviation: PD, Parkinson's disease*

### Differentially expressed genes in COVID-19

To identify differentially expressed genes specific to COVID-19, we analyzed changes to gene expression between COVID-19 and Control groups (see Supplementary Table 1a-g for complete lists for all cells and individual cell types). Different cell types were analyzed individually, and genes primarily involved in responses to immune and viral infections were differentially expressed (Supplementary Figure 3). Top gene candidates were differentially expressed with log2FC>|3| (8-fold change and above) and were significant after correcting for multiple testing (BH p-value<0.05).

Across all cell types, the top COVID-19 associated genes meeting significance thresholds include *SERPINI2*, *ANKRD2*, *FOXJ1*, *RANBP3L*, *FOXB1*, and *IFI44L* (upregulated), and *LIPG*, *ACY3*, *HOXB2*, *MPEG1*, and *LSP1* (downregulated). These genes are broadly associated with interferon-related immune responses, ciliogenesis and cytoskeletal organization, transcriptional regulation, lipid metabolism, metabolic processing, developmental regulation, and immune-associated signaling.

Among the oligodendrocytes, top genes are associated with ion transport (*FXYD6*), synaptic organization (*GPC5*, *LRRC7*), lipid metabolism (*PNPLA3*) and developmental regulation (HOXB2*, GPC5*). In astrocytes, genes reflect functions related to ciliary structure (*DYDC2*, *C90rf135*, *C11orf16*, *ANKRD66*), ion channel activity (*TRPC6, CLIC6*), cytokine signaling (*IL5RA*), immune-associated (*OLR1*, *LSP1*, *CX3CR1*, *ITGAX*) and adhesion processes (*SUSD3*, *EMILIN2*, *CNTNAP2*), and other regulatory processes (*FAM216B*, *BTBD17*, *DDTL*, *FAM180B*, *MAB21L2*). Microglia-associated genes are associated with interferon-related immune response (*IFI44L*, *ISG15*, *IFITM3*, *LY6E*, *SIGLEC1*, *CD163*, *TRIM58*), stress response pathways (*BAG3*, *HSPB1*), lipid and metabolic processes (*LIPG*, *ACSBG1*, *CYP26C1, SGCD*), and neuronal signaling (*GRID2*, *GPR158*). Within OPCs, top genes include transcriptional regulators (*FOXG1*, *HOXB2*), metabolic enzymes (AKR1C3, ATP8B4), and immune-related signaling components (*CX3CR1*, *ITGAX*, *CSF3R*, *P2RY13*, *MILR1*), and developmental transcriptional programs (*WDFY4*). Neuronal cells expressed genes associated with immune signaling (*IFI6*), intracellular trafficking (*KLC3, MYH7*), stress response and protein homeostasis (*MIA*), and receptor-mediated signaling pathways (*ADGRD2*, *INSRR*, *NTRK1*, *EFCAB1*). Within the vascular-associated cluster, gene expression patterns are highlighted by metabolic and enzymatic regulators (*SPR*, *MARS2*), stress-response chaperones (*HSPA1L*), and intracellular trafficking components (*KIF19, RAB7B*).

### Differentially expressed genes in Parkinson’s disease

Differentially expressed genes for PD were identified by pairwise comparison between the PD and Control groups (for complete lists for all cells and individual cell types, see Supplementary Table 2a-g). All cell types were analyzed individually, and genes that are mostly involved in relevant PD pathways including neurotransmitters and dopaminergic activity (Supplementary Figure 4). Top gene candidates were differentially expressed with log2FC>|3| and were significant after correcting for multiple testing (BH p-value<0.05).

Across all cell types, the top genes are broadly associated with neuronal signaling (*GPR6*, *HTR6*, *KCNJ4*, *KCNG3*, *LOXHD1*, *SH3RF2*), transcriptional regulation (*DLX6*, *MYB*, *FOXG1*, *HOXB2*), ion channel activity (*KCNJ4*, *KCNG3*), lipid and broader metabolism (*LIPG*, *LDLR*, *ACY3*), and immune-associated processes (*TESPA1*, *NLRP1*, *ITGAX*, *MYB*, *SUSD3*).

In oligodendrocytes, genes reflect functions related to synaptic organization (*NRGN*, *LRRC7*), neuronal/sensory signaling (*PLCL2*, *OR2L5*, *OR2L3*, *CDH13*, *NEFH*), cell adhesion (*ITGB4*, *CDH13*, *LSR*), and cytoskeletal structure (*FIGN*, *FRY*, *NEFH*), alongside genes involved in lipid and lipoprotein metabolism (*LSR*, *LDLR*, *LIPC*), growth factor signaling (*PDGFRB*), and transcriptional regulation (*HOXB2*). Astrocytes show genes associated with developmental transcriptional regulation, and their cellular differentiation (*FOXG1*, *FEZF2*, *LHX2*, *SIX3*, *NKX6-1*, *CRABP1*, *RET*), GPCR and broader receptor/signaling pathways (*ADGRV1*, *LGR6*, *PRLHR*, *ADRA2A*, *WIF1*, *EPHA6*, *RET*), intracellular trafficking and ubiquitin/protein-turnover processes (*SNX31*, *USP6*), and neurotransmitter or neuronal excitability-related signaling (*GABRD*, *ADRA2A*, *DAO*, *PRLHR*, *ADGRV1*, *EPHA6*), in conjunction with genes involved in glycosphingolipid/cell-surface glycan biology (*A4GALT*) and less-characterized extracellular or ciliary-associated processes (*FAM180B*, *C1orf87*). In microglia, *S100A9, S100A8, DYSF, FAM20A, NRGN, CD163, OCLN, FRY, PPP1R1BM,* and *TUBA1B* were upregulated, while *USP6, PROXC2, and CGNL1* were downregulated. These genes are linked to inflammatory signaling, cytoskeletal organization, membrane dynamics, and transcriptional regulation. Within the OPC cluster, top genes were linked to developmental transcriptional regulation and differentiation (*FOXG1*, *SIX3*, *LHX2*, *HES5*, *VAX1*, *HOXB2*, *PAX3*), signaling regulation (*ADGRV1*, *NELL1*, *CAV1*, *RASAL3*, *USP6, TPD52L1*), cytoskeletal/structural organization (*NEFH*, *FRY*), and synaptic/neuronal signaling (*NRGN*, *NEFH*). Within the neuron cluster, top genes were associated with neuronal signaling/neuropeptide activity (*CRH*, *SLC26A4*), transcriptional regulation and developmental patterning (*DLX2*, *DLX5*, *DLX6*, *NKX2-1*, *LHX6*, *SP8*, *DMBX1*, *SOX14*, *TFAP2B*, *HOXC4*, *HOXA4*, *HOXB2*, *HOXB3*, *LHX5*, *IRX1*, *IRX2*), and extracellular/structural processes (*MEPE*, *HYPM*). In vascular-associated cells, *IL6, TPD52L1, and NSA2* were upregulated, and F*YB1, FAT2, FGD2, DEF6, LAT2, SLC2A5, FBLN7, RHBDF2, GPRIN3,* and *RASGEF1C* were downregulated. Gene signatures involved with cytokine signaling, transcriptional regulation, metabolic processes, and immune-related functions.

### Differentially expressed genes in Parkinson’s disease and COVID-19

To identify differentially expressed genes specific for both PD and COVID-19, we analyzed changes to gene expression between PD/COVID-19 and Control groups (for complete lists for all cells and individual cell types, see Supplementary Table 3a-g). All cell types were analyzed individually, and genes that are mostly involved in the response to viral infection were differentially expressed in these cell types (Figure 3). Top gene candidates noted were differentially expressed with log2FC>|3| and were significant after correcting for multiple testing (BH p-value<0.05).

**Figure 3.**
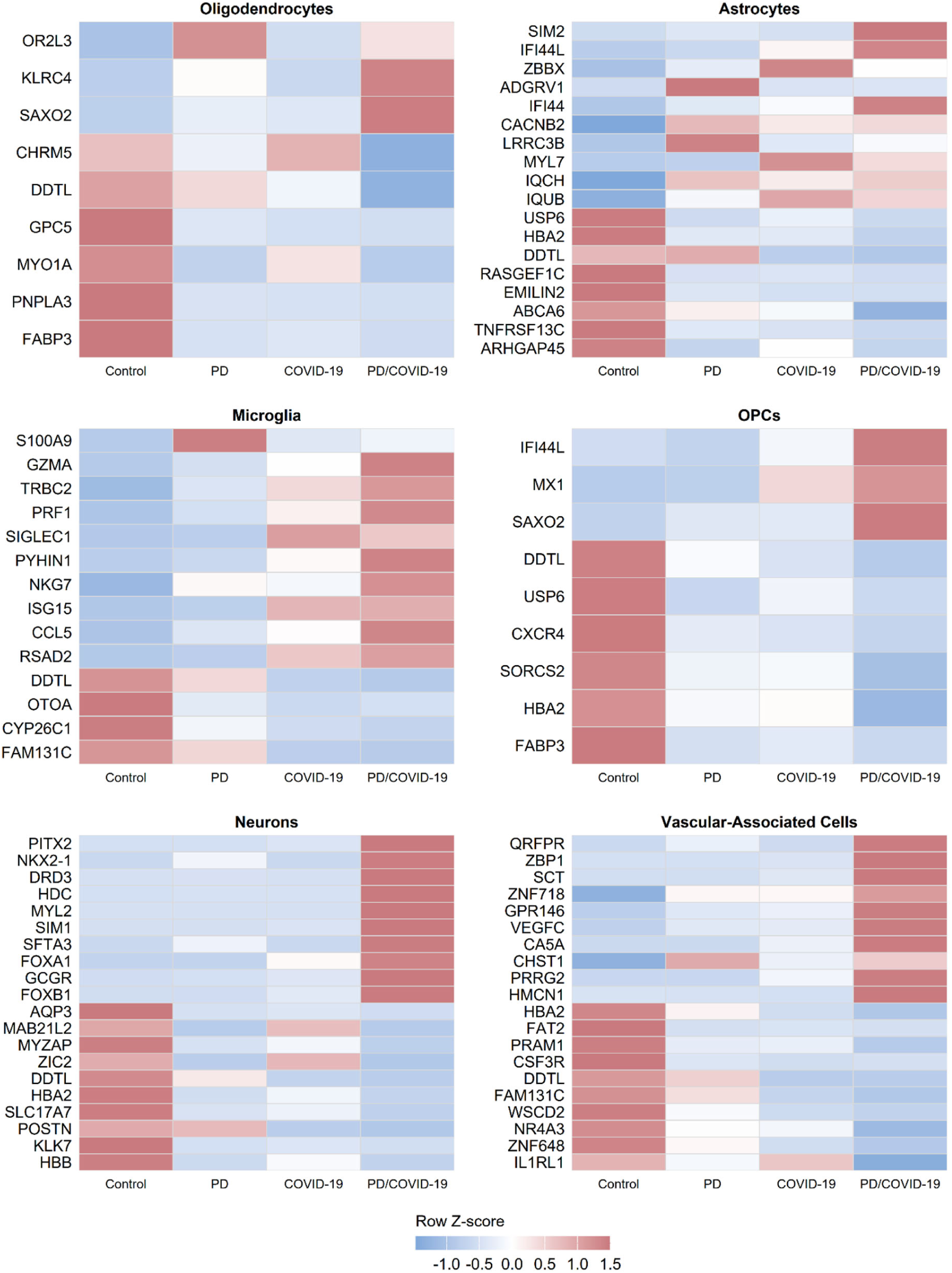
Top differentially expressed Parkinson’s Disease and SARS-CoV-2-associated markers. The top differentially expressed genes (BH-adjusted p < 0.05; |log₂FC| > 3) identified from pairwise comparisons between the PD/COVID-19 and Control groups, are displayed. Heatmap values represent row-scaled (Z-score) mean expression across all four condition groups: Control, PD, COVID-19, and PD/COVID-19; providing cross-condition expression patterns to be assessed for genes defined by their significance within the comorbidity group (PD/COVID-19)

Across all cell types within the PD/COVID-19 group, top genes were linked to interferon/antiviral responses (*IFI44L*, *RSAD2*, *ISG15*), ciliary structure and motility (*DNAAF1*, *DNAH7*, *SAXO2*, *HYDIN*), vesicle/cellular signaling (TPD52L1, SERPINI2, OR2L3), immune-associated signaling (KLRC4), cytoskeletal or cellular organization processes (MYO1A, MEIKIN), and metabolic processes (*DDTL*).

In oligodendrocytes, top genes reflected ciliary structure (*SAXO2*), immune-associated signaling (*KLRC4*), GPCR activity (*OR2L3*, *CHRM5*), lipid/metabolic processes (*PNPLA3*, *FABP3*, *DDTL*), cytoskeletal organization (*MYO1A*), and cellular signaling (*GPC5*). Within the astrocyte cluster, top genes were associated with transcriptional regulation (*SIM2*, *ZBBX*), interferon-related signaling (*IFI44L*, *IFI44*), GPCR and calcium signaling (*ADGRV1*, *CACNB2*, *LRRC3B*, *RASGEF1C*), cytoskeletal processes (*MYL7*, *IQCH*, *IQUB*, *ARHGAP45*), ubiquitin-related signaling/trafficking (*USP6*), metabolic regulation (*HBA2*, *DDT*L, *ABCA6*), extracellular matrix organization (*EMILIN2*), and immune signaling (*TNFRSF13C*). Among microglia, top genes were broadly associated with inflammatory and immune-related signaling (*S100A9*, *SIGLEC1*, *CCL5*, *PYHIN1*), cytotoxic immune processes (*GZMA*, *TRBC2*, *PRF1*, *NKG7*), interferon responses (*ISG15*, *RSAD2*), cell-cycle/ubiquitin-related regulation (*DTL*), retinoid/metabolic processing (CYP26C1), and extracellular/structural organization (*OTOA*, *FAM131C*). Within the OPC cluster, top genes were associated with interferon-regulated signaling (*IFI44L*, *MX1*), ciliary structure and cellular organization (*SAXO2*), chemokine signaling (*CXCR4*), neuronal receptor activity (*SORCS2*), ubiquitin-related signaling/trafficking (*USP6*), and metabolic or oxygen-handling processes (*DDTL*, *FABP3*, *HBA2*).

In neurons, top genes reflected transcriptional regulation and developmental patterning (*PITX2*, *NKX2-1*, *SIM1*, *FOXB1*, *FOXA1*, *MAB21L2*, *ZIC2*), neuronal signaling and neurotransmission (*DRD3*, *HDC*, *SLC17A7*), GPCR/hormonal signaling (*DRD3*, *GCGR*), cytoskeletal or junctional processes (*MYL2*, *MYZAP*), extracellular matrix organization (*POSTN*, *KLK7*, *SFTA3*), and metabolic/transport or oxygen-handling processes (*AQP*3*, DDTL*, *HBA2*, *HBB*). Among vascular-associated cells, top genes were associated with GPCR and neuropeptide/hormonal signaling (QRFPR, SCT, GPR146), transcriptional regulation (ZNF718, NR4A3, ZNF648), angiogenesis and vascular signaling (VEGFC), extracellular matrix/structural organization (HMCN1, FAT2, WSCD2, FAM131C), immune signaling (ZBP1, PRAM1, CSF3R, IL1RL1), metabolic or oxygen-handling processes (CA5A, CHST1, HBA2, DDTL), and membrane/cellular signaling (PRRG2).

### Differentially expressed PD/COVID-19 genes shared with COVID-19 and PD groups that show amplified magnitude

To investigate potential interaction between PD and COVID-19, we next examined genes that were significantly altered in the same direction across PD, COVID-19, and PD/COVID-19 groups. We defined an amplified comorbidity-associated effect as genes for which the absolute log2 fold-change in the PD/COVID-19 group exceeded that observed in either the PD or COVID-19 condition alone (|log2FC|>1). The log2 fold-change is on a transformed scale, therefore, amplification was interpreted as an increase in effect magnitude rather than strict arithmetic additivity. For descriptive purposes, amplification was quantified relative to the stronger of the two single-condition effects.

Using this approach, we identified a subset of genes across multiple cell types that demonstrated increased magnitude of expression changes in the PD/COVID-19 group (Figure 4). Across all cell types, *DDTL* (3.51x) consistently exhibited amplified suppression.

**Figure 4.**
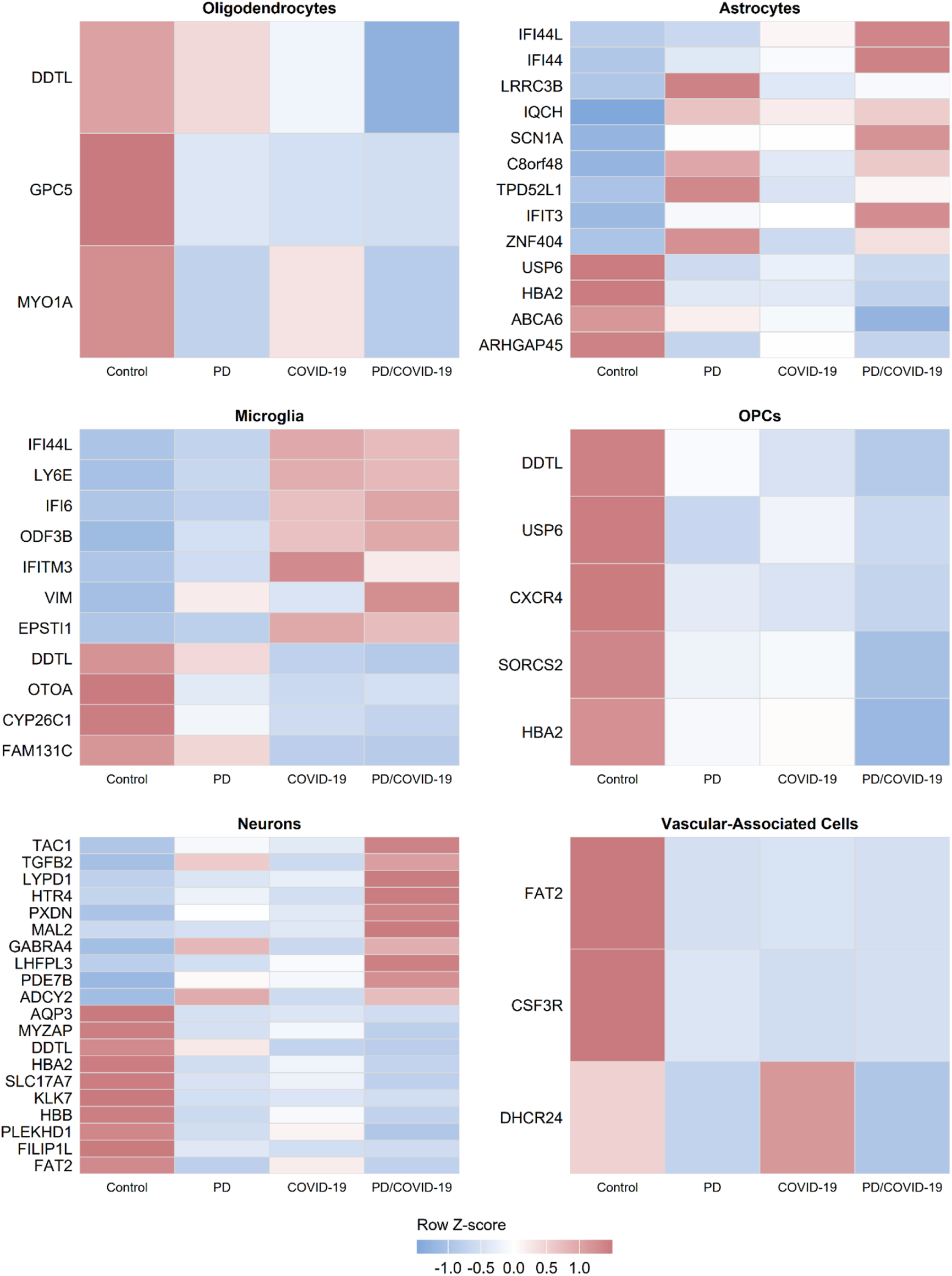
Top differentially expressed genes amplified by PD and COVID-19 Comorbidity. Differentially expressed genes (up to 10 per cell type; BH-adjusted p < 0.05) identified from pairwise comparisons between each group (COVID-19, PD, PD/COVID-19) to control were further analyzed to determine amplified effects within the PD/COVID-19 condition. Amplified comorbidity-associated effect as genes for which the absolute log2 fold-change in the PD/COVID-19 group (|log₂FC|>3) exceeded that observed in either one of the COVID-19 or PD comparisons alone (|log2FC|>1)

Within oligodendrocytes, amplified downregulation affected metabolic/cytoskeletal and cellular signaling genes (*DDTL, MYO1A, GPC5*). In astrocytes, amplified upregulation was enriched for interferon-related genes (*IFI44L, IFI44, IFIT3*), alongside cellular signaling/regulatory genes (*LRRC3B, SCN1A, TPD52L1*), while downregulated genes included lipid/metabolic and cytoskeletal regulators (*ABCA6, DDTL, HBA2, ARHGAP45*). In microglia, amplified upregulation involved interferon and immune-associated genes (*IFI44L, LY6E, IFI6, IFITM3, EPSTI1, VIM*), while downregulated genes included metabolic/retinoid and structural genes (*DDTL, CYP26C1, OTOA, FAM131C*).

Within the neuronal cell cluster, amplified changes involved neuronal signaling and synaptic genes (*TAC1, HTR4, GABRA4, ADCY2, PDE7B*), with downregulation of neurotransmission/metabolic and structural genes (*SLC17A7, DDTL, HBA2, HBB, MYZAP, FAT2*). In OPCs, amplified downregulation affected chemokine/receptor signaling and metabolic genes (*CXCR4, SORCS2, FABP3, DDTL, HBA2*). In vascular-associated cells, amplified downregulation included immune, extracellular matrix, and cholesterol-biosynthesis-associated genes (*CSF3R, FAT2, DHCR24*).

### Pathway Analysis across groups

Gene set enrichment analysis (GSEA) was performed on all cells using KEGG pathways to identify biological processes associated with each condition relative to control (Figure 5). In the COVID-19 group, enrichment was observed in pathways related to receptor-mediated signaling and immune responses, alongside moderate representation of mitochondrial and neurodegeneration-associated gene sets. In contrast, the *PD* group demonstrated enrichment of pathways associated with mitochondrial function and neurodegeneration, including oxidative phosphorylation and Parkinson’s disease-related pathways, consistent with established disease-associated transcriptional changes.

**Figure 5.**
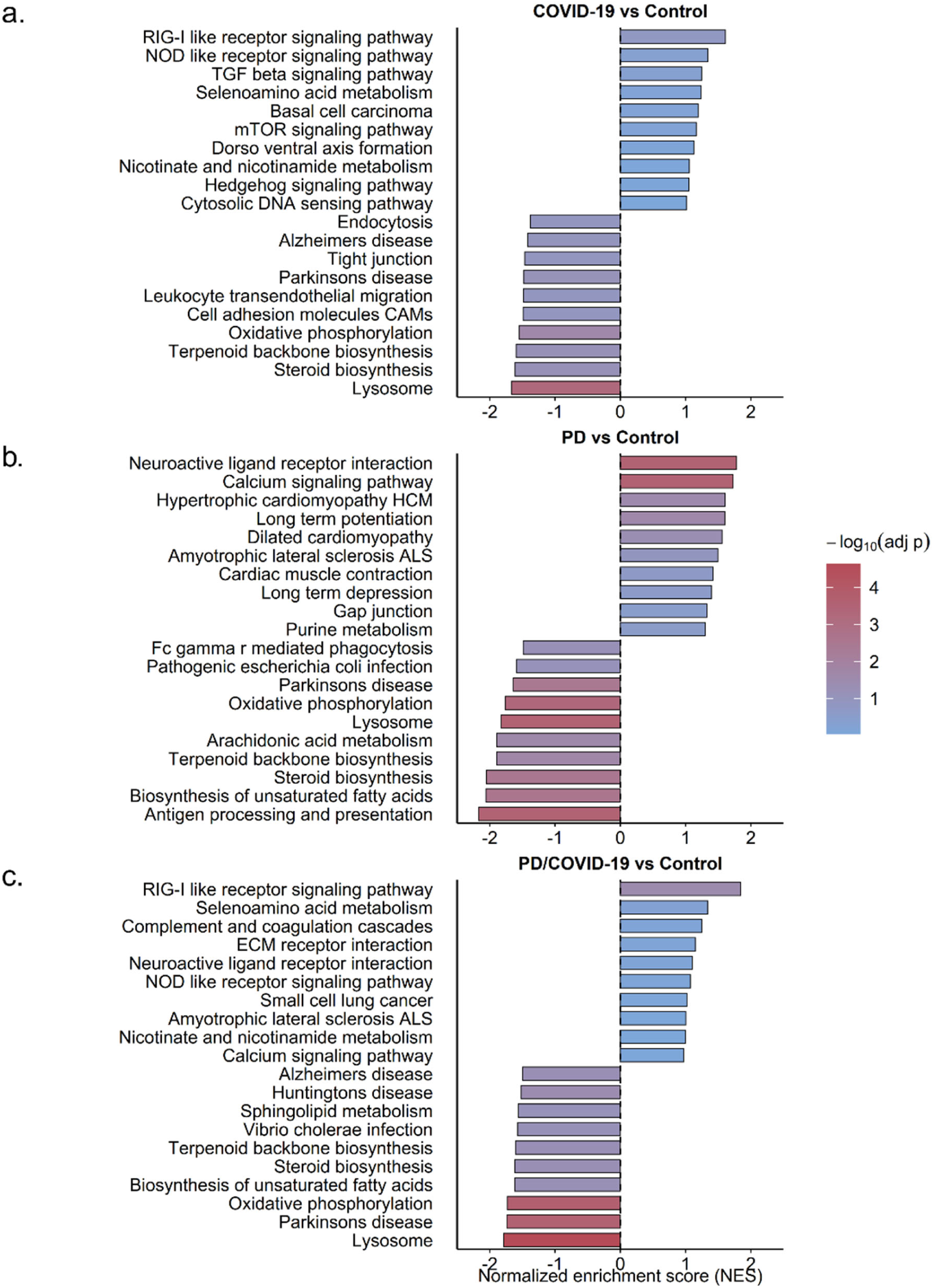
KEGG pathway enrichment across PD, COVID-19, and Comorbid groups. KEGG gene set enrichment analysis was performed on all cells comparing COVID-19 (a), PD (b), and PD/COVID-19 (c) groups relative to Control. The x-axis shows the normalized enrichment score (NES), where positive values indicate enrichment among upregulated genes and negative values indicate enrichment among downregulated genes. Color indicates statistical significance (−log₁₀ adjusted p-value)

The PD/COVID-19 group exhibited a combined enrichment profile characterized by strong upregulation of innate immune and inflammatory signaling pathways, including RIG-I-like receptor signaling, NOD-like receptor signaling, and complement and coagulation cascades. Concurrently, there was coordinated downregulation of pathways associated with mitochondrial function and neuronal homeostasis, including oxidative phosphorylation, lysosomal pathways, and neurodegeneration-associated gene sets (e.g., Parkinson’s disease, Alzheimer’s disease, and Huntington’s disease pathways). This pattern reflects the simultaneous presence of immune activation and suppression of metabolic and neuronal processes within the comorbid condition.

To determine the cellular origin of the pathway-level changes observed across all cells, KEGG GSEA was performed within major cell types for the PD/COVID-19 group relative to control (Supplementary Figure 5).

Microglia exhibited strong enrichment of innate immune and inflammatory signaling pathways, including RIG-I-like receptor signaling, Toll-like receptor signaling, cytosolic DNA sensing, and complement and coagulation cascades. Concurrently, pathways related to lipid metabolism and cellular communication, including glycerophospholipid metabolism and gap junction signaling, were relatively reduced, indicating a shift toward an activated immune state.

Astrocytes similarly demonstrated enrichment of immune-associated pathways, including RIG-I-like receptor signaling and antigen processing and presentation, alongside modulation of metabolic and signaling pathways such as mTOR signaling. In parallel, astrocytes showed reduced enrichment of mitochondrial and lysosomal pathways, including oxidative phosphorylation and lysosome-associated processes, consistent with altered cellular homeostasis.

In contrast, neurons exhibited enrichment of pathways related to cellular signaling and structural organization, including neuroactive ligand–receptor interaction, calcium signaling, and axon guidance. At the same time, pathways associated with metabolic processes, including steroid biosynthesis, were relatively reduced, indicating alterations in neuronal signaling and homeostatic function.

### Convergence of global Proteome with cell-specific transcriptome to highlight infection related signature

To validate the pathways identified by single-nucleus RNA sequencing (snRNA-seq), we performed global proteomic profiling of substantia nigra tissue derived from the same samples used for the transcriptomic analyses.

In total, 4,532 proteins were detected across all samples, with modest differences in protein coverage between groups (Supplementary Figure 6a, Supplementary Table 4a). The Control group contained 3,681 detected proteins, whereas the PD group contained 4,206 proteins, the COVID-19 group contained 4,192 proteins, and the PD/COVID-19 group contained 3,830 proteins. Despite these group-specific differences, a large conserved core proteome was observed, with approximately 3,351 proteins shared across all experimental groups.

A smaller subset of proteins displayed condition-specific detection patterns. Specifically, 46 proteins were exclusively detected in the Control group, 187 proteins were unique to the PD group, 89 proteins were uniquely detected in the COVID-19 group, and 18 proteins were exclusively identified in the PD/COVID-19 group. Notably, 274 proteins were shared among all disease-associated groups but were absent in controls (Supplementary Figure 6b, 6c; Supplementary Table 4b).

Principal component analysis (PCA) of the proteomic dataset, with principal component 1 (PC1) explaining 31% of the variance and principal component 2 (PC2) explaining 11% of the variance, demonstrated clear clustering of Control samples, while PD samples also formed a distinct cluster. The PD/COVID-19 group exhibited particularly tight clustering, indicating a comparatively homogeneous proteomic profile within the comorbid condition. In contrast, COVID-19 samples displayed greater dispersion, consistent with increased inter-individual variability (Figure 6a). Intergroup relationships were further supported by correlation analysis, which demonstrated increased similarity between the COVID-19 and PD/COVID-19 groups (Figure 6b). Likewise, hierarchical clustering heatmaps revealed close clustering of the COVID-19 and PD/COVID-19 proteomic profiles (Figure 6c).

**Figure 6.**
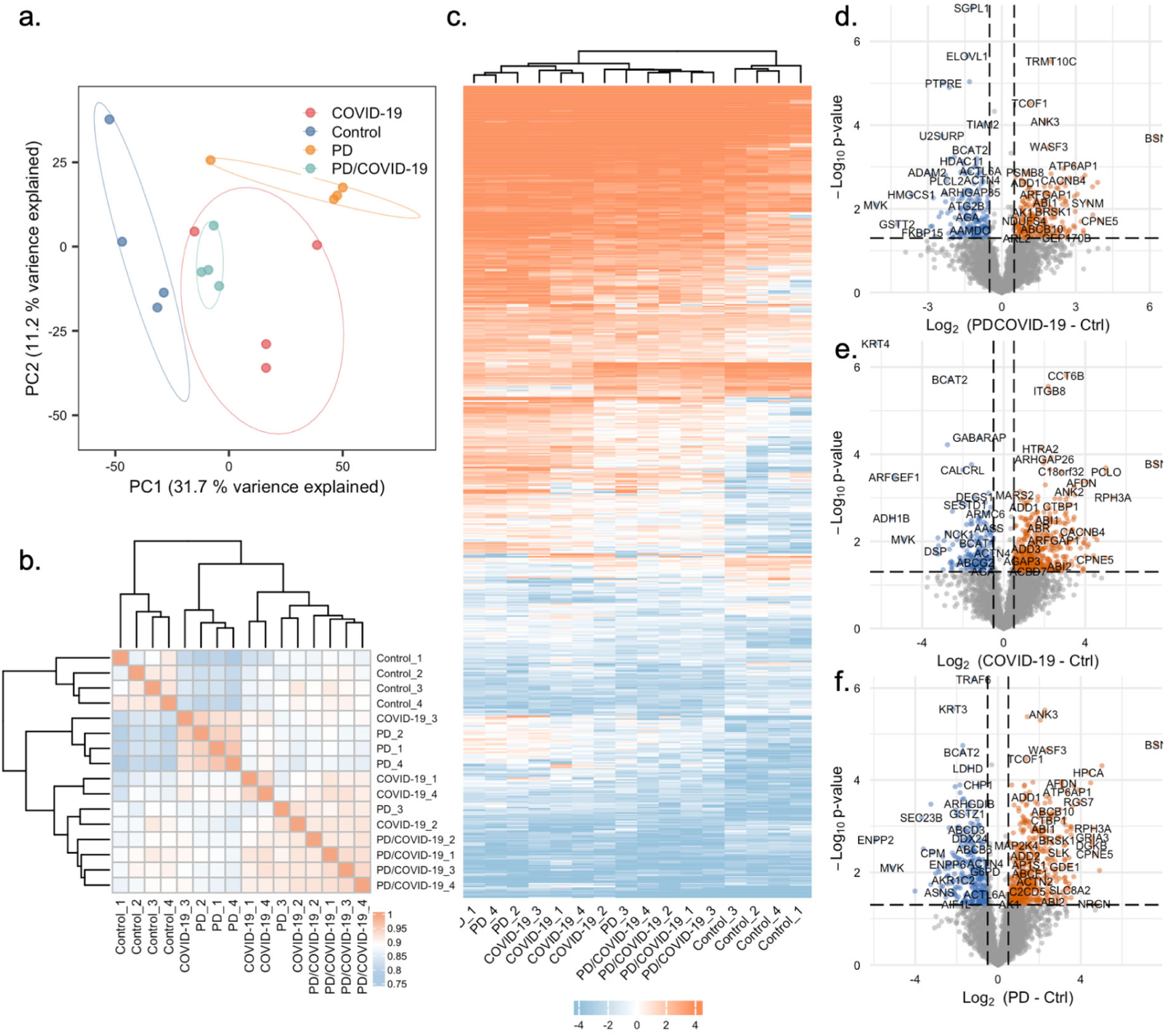
Global proteomic profiling of substantia nigra tissue across Parkinson’s disease (PD), COVID-19 (Covid-19), comorbid (PD/COVID-19) group and controls (Control). Principal component analysis (PCA) of proteomic profiles across samples (a). Each point represents an individual sample, projected onto the first two principal components (PC1 and PC2), which capture the largest sources of variance in the dataset. Clustering of samples reflects similarity in protein expression patterns, while separation between groups (Control, PD, COVID-19, PD/COVID-19) indicates condition-specific proteomic differences. Correlation matrix of samples based on protein abundance profiles (b). Pearson’s correlation coefficients were computed using pairwise complete observations. Samples are hierarchically clustered, and group annotations (Control, PD, COVID-19, PD/COVID-19) are indicated. Warmer colors indicate higher similarity between samples. Heatmap of row-scaled protein abundance across all samples (Control, PD, COVID-19, PD/COVID-19; (c)). Proteins were z-score normalized across rows. Columns are hierarchically clustered, while rows are either clustered. The color scale represents relative abundance (blue: low, white: zero, red: high). Volcano plots comparing PD/COVID-19, COVID-19, and PD proteome with that of control samples (d-f). Significant proteins reflect combined or interaction effects of PD and COVID-19 conditions. The x-axis represents log2 fold change, and the y-axis represents −log10 p-values. Proteins passing significance thresholds (log2FC > 0.5, or log2FC > 0.5, and p < 0.05) are highlighted (orange: upregulated; blue: downregulated). Non-significant proteins are shown in grey

### Differential protein abundance analysis

To further assess biologically relevant alterations in protein abundance, differential expression analyses were performed using a significance threshold of P≤0.05 together with a log2 fold-change cutoff of >0.5 for upregulated proteins and <−0.5 for downregulated proteins. Pairwise comparisons were conducted between PD versus Control, COVID-19 versus Control, and PD/COVID-19 versus Control groups. The resulting volcano plots are shown in Figure 6d-f (Supplementary Table 4c). Applying these thresholds, 288 proteins were identified as downregulated and 454 proteins as upregulated in the PD group compared with Controls, whereas 3,790 proteins were not differentially regulated. The top 10 upregulated candidates, shown as protein name (+log2FC), included BSN (+7.66), PCLO (+5.02), PDE10A (+4.91), CPNE5 (+4.68), NRGN (+4.66), RPH3A (+4.59), GRIA3 (+4.56), SYNGAP1 (+4.52), DGKB (+4.52), and PPP1R12A (+4.48). The top 10 downregulated candidates, shown as protein name (−log2FC), included ENPP2 (−5.91), MVK (−5.12), ZDHHC14 (−4.02), SEC23B (−3.70), TRIM36 (−3.61), GATAD2A (−3.25), CASP14 (−3.24), CPM (−3.14), DSP (−3.02), and ITGA6 (−2.89).

Similarly, in the COVID-19 group, 170 proteins were downregulated and 414 proteins were upregulated relative to the controls, while 3,948 proteins remained unchanged. The top 10 upregulated candidates included BSN (+7.485), RPH3A (+5.36), PCLO (+5.03), DGKB (+5.02), ZC3HAV1 (+5.0), PTPRN (+4.58), CPNE5 (+4.50), HPCA (+4.46), GAD2 (+4.41), and SCG2 (+4.29). The top 10 downregulated candidates included KRT4 (−6.27), ADH1B (−5.50), ARFGEF1 (−5.35), MVK (−4.924), DSP (−3.338), LSS (−3.225), TMEM97 (−3.224), KLK6 (−3.192), DOCK5 (−2.968), and PLBD2 (−2.925).

In the PD/COVID-19 group, 184 proteins were downregulated and 223 proteins were upregulated compared with controls, whereas 4,125 proteins were not differentially regulated. The top 10 upregulated candidates included BSN (+6.274), CPNE5 (+3.976), PCLO (+3.884), HPCA (+3.694), RGS7 (+3.680), SYNM (+3.488), DDRGK1 (+3.373), GAD2 (+3.320), ZC3H15 (+3.304), and RPH3A (+3.267). The top 10 downregulated candidates included MVK (−5.120), GSTT2 (−4.253), HMGCS1 (−3.672), FKBP15 (−3.227), ADAM2 (−3.024), RPL35A (−2.890), PPIL3 (−2.837), UBLCP1 (−2.826), CALB1 (−2.785), and ASNS (−2.719) (Supplementary Table 4c).

### Pathway enrichment analysis of global proteomics across the groups

To determine whether the proteomic alterations observed across the disease groups converged on specific biological processes, we performed Gene Ontology (GO) Biological Process enrichment analysis using ranked protein abundance changes relative to Control groups.

In the PD group, the most significantly enriched upregulated pathways were predominantly associated with synaptic function and neuronal communication. These included neurotransmitter secretion, neurotransmitter transport, vesicle-mediated transport in synapse, synapse organization, trans-synaptic signaling, cell-cell signaling, and cell junction organization (Figure 7a, Supplementary Table 5a). In contrast, downregulated pathways were primarily related to lipid and sterol homeostasis, including sterol metabolic process, steroid biosynthetic process, alcohol metabolic process, sphingolipid metabolic process, and steroid metabolic process. Additional suppressed pathways included tumor necrosis factor response, amide metabolic process, and keratinization (Figure 7a, Supplementary Table 5a).

**Figure 7.**
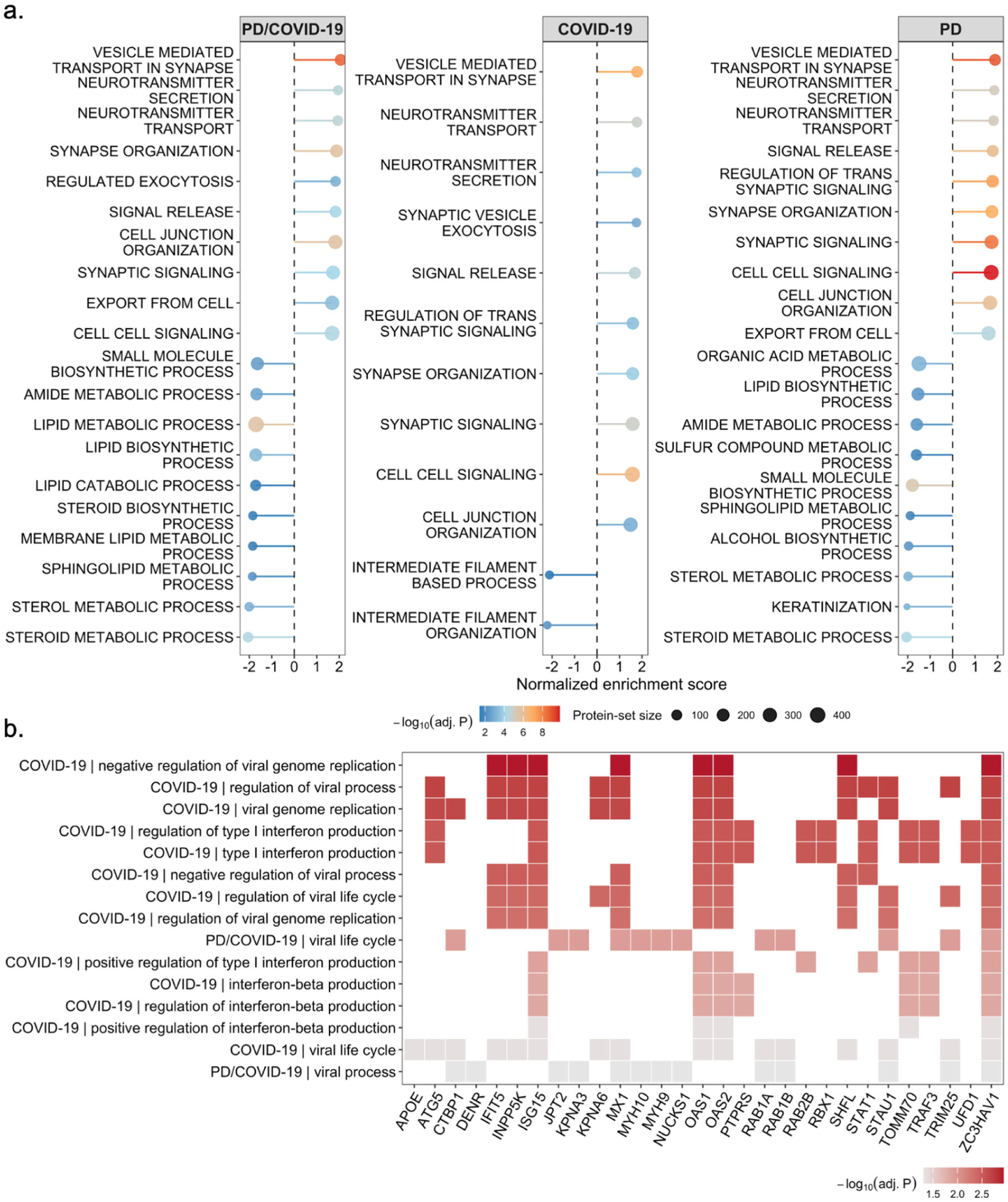
Viral response pathways enriched in COVID-19-associated proteomic signatures - Gene Ontology pathway alterations associated with proteomic changes in Parkinson’s disease, Covid-19, and their comorbidity. Gene Set Enrichment Analysis of Gene Ontology Biological Process terms was performed using ranked proteomic changes in PD, Covid-19, and PD/COVID-19 groups compared with Control controls (a). Significantly enriched pathways are shown according to normalized enrichment score. Dot size indicates the number of proteins associated with each pathway, and dot color represents the adjusted *p*-values. A heatplot showing Over-Representation Analysis of significantly upregulated and down-regulated proteins identified in the COVID-19 and PD/COVID-19 groups relative to controls (b). Enriched pathways, including viral process, viral life cycle, viral genome replication, regulation of viral genome replication, regulation of viral process, and negative regulation of viral genome replication were exclusively found to be associated with COVID-19 and PD/COVID-19 upregulated proteins. Color indicates the adjusted *p*-value for pathway enrichment

Interestingly, the proteomic pathway signature identified in the COVID-19 group closely resembled that observed in the PD cohort. Similar enrichment of neuronal and synaptic processes was detected, including neurotransmitter secretion, neurotransmitter transport, vesicle-mediated transport, trans-synaptic signaling, regulation of signal release, cell-cell communication, and cell junction organization. Conversely, pathways associated with intermediate filament organization, sterol and steroid metabolic processes, keratinization, epidermis development, and lipid metabolism were significantly reduced relative to controls (Figure 7a, Supplementary Table 5b).

Analysis of the PD/COVID-19 group revealed a largely overlapping molecular signature. Upregulated biological processes were again dominated by pathways involved in neurotransmission, synaptic organization, vesicle trafficking, and neuronal signaling. Downregulated pathways included sterol metabolic process, steroid biosynthetic process, membrane lipid metabolic process, androgen metabolic process, amide metabolic process, and small-molecule biosynthetic pathways (Figure 7a, Supplementary Table 5c).

Because ranked enrichment analyses are often dominated by broad biological trends, and proteins uniquely detected in individual disease groups are not captured by GSEA, we next sought to identify disease-specific inflammatory and infection-associated signatures by performing over-representation analysis (ORA) separately on significantly upregulated (along with uniquely identified proteins in respective groups e.g., PD/COVID-19 upregulated proteins and PD/COVID-19 uniquely identified proteins together) and downregulated proteins using the same Gene Ontology Biological Process database. This approach enabled the identification of discrete biological pathways enriched among the most strongly altered proteins and allowed more focused assessment of immune and infection-related signaling events associated with Parkinson’s disease, COVID-19, and their comorbidity.

The over-representation analysis (ORA) largely corroborated the findings obtained from the GSEA, highlighting a consistent biological signature across the PD, COVID-19, and PD/COVID-19 groups. Among the significantly upregulated proteins, the most enriched biological processes were associated with synaptic function, including synaptic vesicle cycle, vesicle-mediated transport in synapse, neurotransmitter secretion, regulation of signal release, and related neuronal communication pathways. In addition, proteins involved in cytokinesis-associated processes were enriched among the upregulated candidates in both the PD and COVID-19 groups. Conversely, downregulated proteins were predominantly enriched for pathways related to cholesterol, sterol, steroid, and alcohol metabolic processes. A subset of downregulated proteins was also associated with cytokinesis-related pathways (Supplementary Figure 7, Supplementary data 6a-6f).

Importantly, the ORA provided additional resolution of infection-associated biological processes that were not prominent in the ranked GSEA analyses. Comparative cluster enrichment analysis revealed significant enrichment of pathways related to viral life cycle, viral process, host response to virus, and viral genome replication, specifically among the upregulated proteins in the COVID-19 and PD/COVID-19 groups (Figure 7b, Supplementary Table 6c-6e). These enriched categories included GO terms such as negative regulation of viral genome replication, regulation of viral process, viral genome replication, regulation of type I interferon production, type I interferon production, negative regulation of viral process, regulation of viral life cycle, regulation of viral genome replication, positive regulation of type I interferon production, interferon-beta production, regulation of interferon-beta production, positive regulation of interferon-beta production, viral life cycle, and viral process (GO:0045071, GO:0050792, GO:0019079, GO:0032479, GO:0032606, GO:0048525, GO:1903900, GO:0045069, GO:0032481, GO:0032608, GO:0032648, GO:0032728, GO:0019058, and GO:0016032). Proteins contributing to these enriched antiviral and interferon-associated pathways included APOE, ATG5, CTBP1, DENR, IFIT5, INPP5K, ISG15, JPT2, KPNA3, KPNA6, MX1, MYH9, MYH10, NUCKS1, OAS1, OAS2, PTPRS, RAB1A, RAB1B, RAB2B, RBX1, SHFL, STAT1, STAU1, TOMM70, TRAF3, TRIM25, UFD1, and ZC3HAV1.

### Protein candidates overlapping top differentially regulated genes from single-nuclear transcriptome

Next, we examined the abundance profiles of proteins corresponding to the top 55 candidate genes identified by the snRNA-seq analysis. Among these candidates, only ADCY2, DHCR24, GABRA4, HBB, IFIT3, IFITM3, MAL2, SCN1A, SLC17A7, SORCS2, TPD52L1, and VIM were detected within the proteomic dataset (Supplementary Figure 8). Pairwise comparisons for these proteins were performed between the PD/COVID-19 group and the Control, PD, and COVID-19 groups.

Among these candidates, DHCR24 abundance was significantly decreased in the PD/COVID-19 group compared with the Control group. However, DHCR24 levels in PD/COVID-19 samples did not differ significantly from those observed in the PD or COVID-19 groups. In contrast, HBB abundance in the PD/COVID-19 group was significantly increased compared with the PD group, while no significant differences were observed relative to the Control or COVID-19 groups.

No significant differences were detected in the abundance of ADCY2, IFITM3, MAL2, SLC17A7, TPD52L1, or VIM when comparing the PD/COVID-19 group with the other experimental groups. Notably, IFIT3 was detected exclusively in the COVID-19 and PD/COVID-19 groups, although no significant difference in abundance was observed between these two conditions.

Several proteins also exhibited highly restricted detection patterns. GABRA4 was detected exclusively in the PD group, SCN1A was detected only in the PD and Covid-19 groups, and SORCS2 was uniquely detected in the COVID-19 group.

### Amplified proteomic alterations associated with PD/COVID-19 comorbidity

To investigate potential interactions between PD and COVID-19, we next examined proteins that were significantly altered in the same direction across the PD, COVID-19, and PD/COVID-19 groups. Proteins were classified as exhibiting an amplified comorbidity-associated effect when the absolute log2 fold-change observed in the PD/COVID-19 group exceeded that detected in either individual condition alone (|log2FC|>0.58). For descriptive purposes, amplification was assessed relative to the larger of the two single-condition effects.

Using these criteria, we identified a subset of proteins displaying progressively greater abundance changes in the PD/COVID-19 group compared with both PD and COVID-19 groups (Supplementary Figure 9). Among these were ABCD3, ACTN4, CAVIN1, CAVIN2, COL4A2, DDRGK1, DTD1, GSTZ1, NAA15, NAA50, NEMF, SESTD1, SGPL1, SNX17, STS, and WASF2, all of which exhibited larger effect sizes in the PD group than in the COVID-19 group, with a further increase in the comorbid PD/COVID-19 condition.

A second group of amplified proteins included CRTAC1, HDAC6, MYO1C, RUFY2, RUFY3, and TRIM25. In contrast to the proteins described above, these candidates exhibited larger effect sizes in the COVID-19 group than in the PD group, followed by further amplification in the PD/COVID-19 condition.

### Literature Review

The initial PubMed search was designed to identify human studies related to PD, COVID-19, and PD– COVID-19, and brain or post-mortem/CNS contexts, while excluding reviews, meta-analyses, editorials, comments, letters and non-original data (Supplementary Figure 10; Supplementary Tables 7a-f). The PD–COVID-19 query returned 63 records after applying the PubMed human filter.

All records were then manually screened because several retrieved studies did not meet the intended scope of the search. During this first screening step, 35 records were excluded due to not using original data in the study design, using non-human models (11), or using non-brain tissue (1). This resulted in 16 studies retained for further assessment.

There were no studies employing single-cell or single-nucleus RNA sequencing in human brain or central nervous system tissue directly examining the combined effects of PD and SARS-CoV-2 infection.

Five of the remaining studies (Supplementary Table 7f) reported relevant molecular or gene expression–related findings, including differential gene expression, protein aggregation dynamics, and neuroinflammatory signaling pathways associated with PD and/or COVID-19. Among these, two studies employed broader transcriptomic approaches; one utilizing spatial transcriptomics in human brain tissue and another using single-cell RNA sequencing in cerebrospinal fluid, while an additional study reported targeted gene-expression changes related to inflammasome activation. These studies provide indirect evidence of overlapping biological mechanisms such as immune activation, mitochondrial dysfunction, and α-synuclein-associated pathology; however, they lack the cell-type-specific resolution required to delineate these processes at the single-cell level.

Differentially expressed genes identified in the present study overlap with pathways reported in prior PD–COVID-19 literature, including interferon-stimulated genes (*ISG15, IFI44L, RSAD2,* and *MX1*) [11, 64], inflammatory mediators (*IL6* and *TNF*) [29], and inflammasome-related genes (*NLRP3* and *IL1B*) [2]. Additionally, perturbations in α-synuclein biology (*SNCA*) have been observed in experimental models of SARS-CoV-2 infection [28].

## Discussion

In this study, we sought to identify shared and distinct molecular mechanisms in PD and severe SARS-CoV-2 infection. To address this question, we analyzed post-mortem striatal tissue from individuals with PD and concomitant COVID-19, PD without COVID-19, COVID-19 without neurological disease, and unaffected controls. All cases underwent detailed neuropathological characterization, and molecular alterations were investigated using single-nucleus RNA sequencing and complementary proteomic analyses.

Previous neuropathological studies have demonstrated that SARS-CoV-2 infection induces neuroimmune activation characterized by astrocytosis, microgliosis, inflammatory signaling, and disturbances in neuronal homeostasis [32, 36, 43, 49, 50, 53, 59]. Although studies in mice [28], singular case reports [14] and clinical and epidemiological observations in humans point towards a relationship between viral infection and PD [17, 24, 33, 51], and the molecular mechanisms linking severe viral infection to neurodegenerative processes are not fully understood [24]. Our data provide evidence that a major point of convergence between COVID-19 and PD is not direct viral injury, but rather activation of innate immune pathways that overlap with molecular programs already implicated in PD pathogenesis.

A central finding of this study is the robust interferon-associated transcriptional signature observed across multiple cell types in the COVID-19 and PD/COVID-19 groups. Canonical interferon-stimulated genes, including *IFI44L*, *IFI44*, *ISG15*, *RSAD2*, *MX1*, *IFIT3*, and *IFITM3*, were consistently upregulated, particularly within microglia and astrocytes. These findings are consistent with previous reports demonstrating activation of type I interferon pathways in the central nervous system following viral infection [26, 43, 45, 50, 60]. However, the amplified expression of these genes in the PD/COVID-19 group suggests that the coexistence of PD and SARS-CoV-2 infection is associated with enhanced or prolonged activation of antiviral signaling pathways rather than a simple additive effect of the two conditions. At the transcriptomic level, these changes were part of a broader, cell-type-specific response encompassing immune and interferon-associated signaling, neuronal function, metabolic homeostasis, and cellular regulatory processes.

Importantly, the transcriptomic findings were independently supported by proteomic analyses. Although overlap between differentially expressed transcripts and proteins at the individual gene level was limited, which is expected given the distinct biological layers captured by transcriptomics and proteomics, both datasets converged on similar biological processes. Thus, the principal agreement between the two datasets occurring at pathway level, strengthens the biological relevance of the observed signatures. Proteomic analyses identified enrichment of antiviral and host-defense pathways involving proteins such as ISG15, MX1, TRIM25, STAT1, ZC3HAV1, KPNA3, and KPNA6, supporting the presence of a sustained antiviral molecular program beyond the transcriptional level [25, 31, 42]. Notably, TRIM25 was also identified within the viral and antiviral response signatures revealed by the over-representation analysis, suggesting a potential link between persistent antiviral responses and PD–COVID-19 comorbidity. The antiviral/host-defense signature (identified in the gene ontology terms) was more pronounced however in the COVID-19 group compared to the PD/COVID-19 comorbid group at the proteomics level, which could be indicative of the delay in the proteome adaptation against the COVID-19 infection with preexistent PD in comparison to COVID-19 patients without the comorbid Parkinson’s pathology.

The strongest evidence for disease convergence emerged within glial populations, particularly microglia. Microglia from PD/COVID-19 cases displayed increased expression of interferon-responsive genes alongside enrichment of pathways related to RIG-I-like receptor signaling, Toll-like receptor signaling, NOD-like receptor signaling, cytosolic DNA sensing, and complement activation. These pathways are central components of innate antiviral immunity but are also increasingly recognized as contributors to neurodegenerative disease progression [58]. Chronic microglial activation is a well-established feature of PD and has been implicated in dopaminergic neuronal loss through the production of inflammatory cytokines, reactive oxygen species, and alterations in synaptic maintenance [20, 23, 35, 52]. The overlap between viral-response pathways and established PD-associated inflammatory mechanisms suggests that severe viral infection may amplify pre-existing neuroinflammatory states, thereby increasing neuronal vulnerability [13, 20].

Beyond immune activation, a second major theme emerging from both transcriptomic and proteomic analyses was dysregulation of lipid and sterol metabolism. Genes and proteins involved in cholesterol biosynthesis and lipid homeostasis, including LDLR, DHCR24, MVK, and HMGCS1, were consistently reduced across disease groups [34, 41]. At the pathway level, in both single-nucleus RNA sequencing and proteomic analyses, we found suppression of sterol, cholesterol, and lipid metabolic processes. This observation is particularly relevant given growing evidence linking lipid dysregulation, lysosomal dysfunction, and impaired membrane homeostasis to PD pathogenesis [6, 15, 62, 63]. The convergence of PD and COVID-19 on these pathways suggests that disruption of cellular metabolic homeostasis may represent a common downstream consequence of both neurodegeneration and severe viral infection.

Interestingly, proteomic analyses revealed substantial overlap among PD, COVID-19, and PD/COVID-19 groups, with all conditions showing enrichment of pathways associated with synaptic organization, neurotransmitter release, and vesicle trafficking. Similar responses are seen in a wide range of neurological diseases and may reflect compensatory responses to neuronal stress and altered network activity [4]. At the same time, the coexistence of enhanced immune activation and impaired metabolic homeostasis in the PD/COVID-19 group suggests that glial and neuronal responses are tightly interconnected. Thus, immune activation may contribute to metabolic dysfunction and disturbed lipid homeostasis which has been implicated in impaired neuronal resilience and synaptic maintenance in neurodegenerative diseases [57].

Taken together, our findings support a model in which severe viral infection induces molecular programs that partially overlap with pathways involved in PD pathogenesis. Our data suggest that viral infection can establish or amplify a persistent innate immune state characterized by interferon signaling, microglial activation, and metabolic dysfunction, closely resembling signatures observed in immunosenescence [18]. This may therefore act as a disease modifier in susceptible individuals. Observed signatures may not be unique to SARS-CoV-2 but could represent shared molecular responses to severe viral infections, providing a potential mechanistic framework linking these to neurodegenerative diseases.

Our study has several limitations. Due to the complexity of the analyses and the availability of well-characterized post-mortem samples the cohort size is rather limited. Some cellular populations are disproportionately represented. Nevertheless, the convergence of pathways identified from transcriptomic and proteomic analyses supports the robustness of our findings.

Consistent with our literature search, and to our knowledge, no previous study has directly examined the combined effects of PD and SARS-CoV-2 infection in human brain or central nervous system tissue using single-cell or single-nucleus transcriptomic analyses. Data on single-cell and single-nucleus transcriptomic studies examining the interaction between PD and SARS-CoV-2 infection in human brain tissue are rare, and data combining transcriptomic and proteomic datasets are not available, thus, the present study provides an initial high-resolution molecular framework for understanding how severe viral infection intersects with PD. Future studies incorporating larger cohorts and additional neurodegenerative disorders will be important to determine whether the molecular signatures identified here represent a generalizable mechanism linking viral infection to neurodegenerative vulnerability.

## Supporting information

Supplementary Table 1_Differentially_Expressed_Genes_COVID-19_vs_CON

Supplementary Table 2_Differentially_Expressed_Genes_PD_vs_Control

Supplementary Table 3_Differentially_Expressed_Genes_PDCOVID-19_vs_Control

Supplementary Table 4_Differentially_Abundant_Proteins

Supplementary Table 5. Proteomics_GSEA_Results

Supplementary Table 6_Proteomics_GO_Results

Supplementary Table 7_Literature Search

## Data Availability

The mass spectrometry proteomics data have been deposited to the ProteomeXchange Consortium via the PRIDE (partner repository with the dataset identifier PXD080847). All additional data produced in the present study are available upon reasonable request to the authors.

## Acknowledgements and competing interests

We thank the Core Facility Mass Spectrometric Proteomics as part of the Technology Platform Mass Spectrometry (TPMS) at University of Hamburg (UHH) and University Medical Center Hamburg-Eppendorf (UKE) for support with mass spectrometric measurements and analysis funded by the Deutsche Forschungsgemeinschaft (DFG, German Research Foundation) – 518551069. This work was funded by the research consortium NATON. The NATON project (grant no. 01KX2524 and 01KX2121) is part of the National Network of University Medicine, funded by the Federal Ministry of Research, Technology and formerly by the Federal Ministry of Education and Research, Germany. Additional support was provided through a DFG Heisenberg Professorship (grant no. TR 1714/8-1) awarded to Prof. J. Trinh. This work was further supported by the Deutsche Forschungsgemeinschaft (DFG, German Research Foundation) through grant no. TR 1714/5-1 awarded to Professors J. Trinh, M. Glatzel, and C. Klein.

