## Supplementary_Figures for "Convergent Innate Immune and Metabolic Signatures in Parkinson’s Disease and Viral Infection"

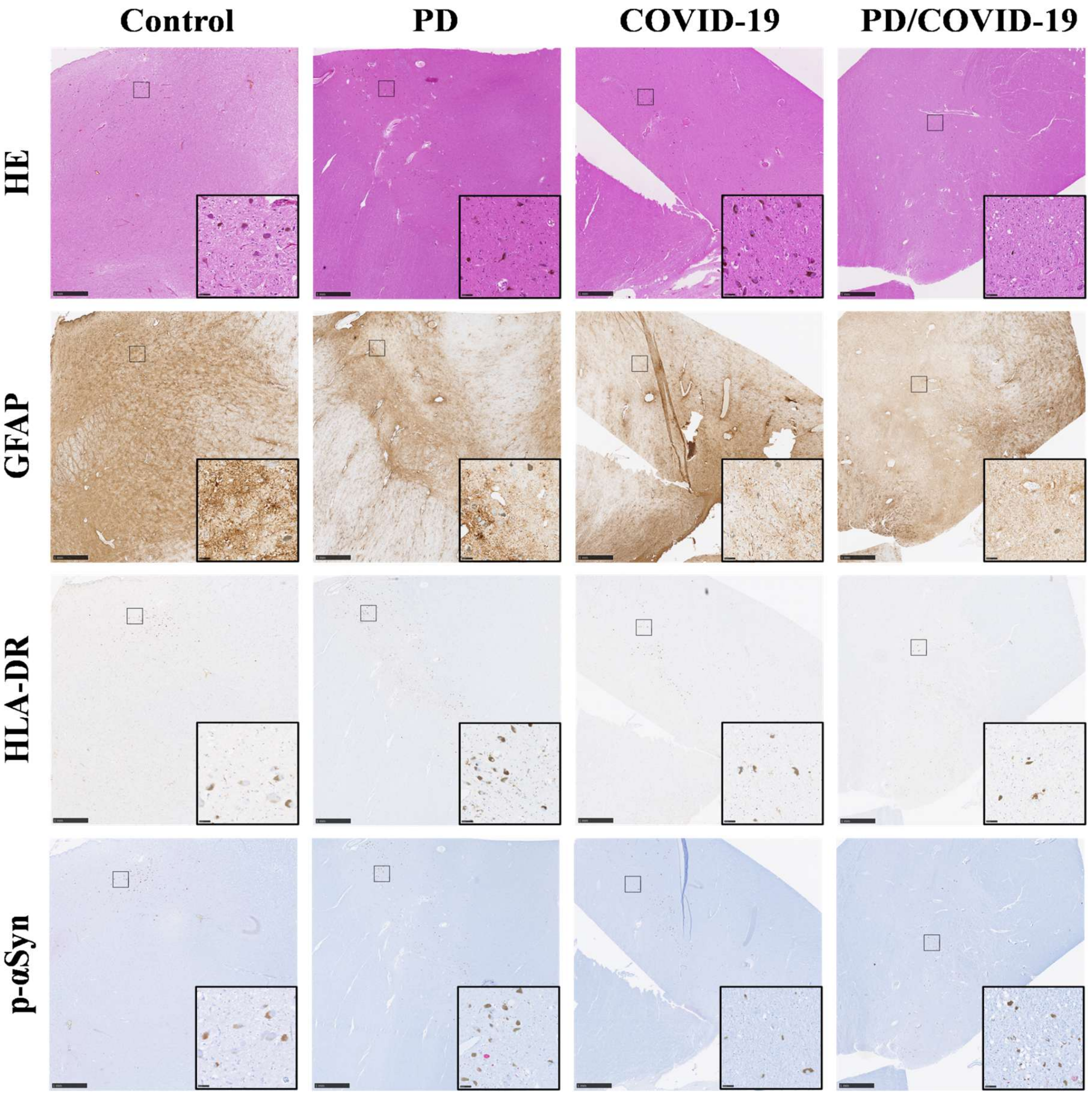


**Supplementary Fig. 1 Histological findings of the substantial nigra of patients with PD/COVID-19, COVID-19, PD, and Control.** Representative images are shown. Haematoxylin and eosin (H&E) staining is shown in the first row. Immunohistochemical staining for the astrocytic marker GFAP (2nd row) demonstrated reactive astrogliosis in PD/COVID-19, COVID-19, PD and control groups. Immunohistochemical staining for the microglial marker HLA-DR (3rd row) showed microglial activation in PD/COVID-19, COVID-19 and PD cases, no activation in controls. Immunohistochemical staining for phosphorylated α-synuclein (p-αSyn) (4th row) showed α-synuclein aggregation in PD/COVID-19, PD cases. Scale bars represent 1 mm (50 µm in inset images)

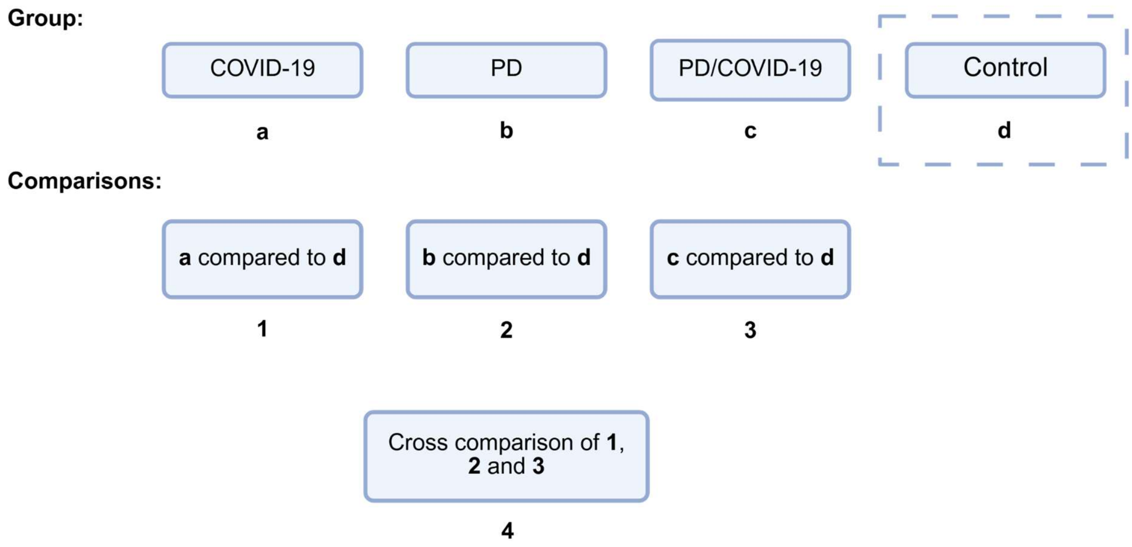


**Supplementary Fig. 2 Schematic overview of the comparisons used for differential expression analyses.** Differential expression was assessed using three pairwise comparisons, with each disease condition (COVID-19, PD, and PD/COVID-19) compared with the control group using the Wilcoxon rank-sum test. To investigate whether transcriptional changes were amplified in the comorbid PD/COVID-19 condition, differentially expressed genes identified across the three comparisons were evaluated for concordance in direction and differences in effect magnitude

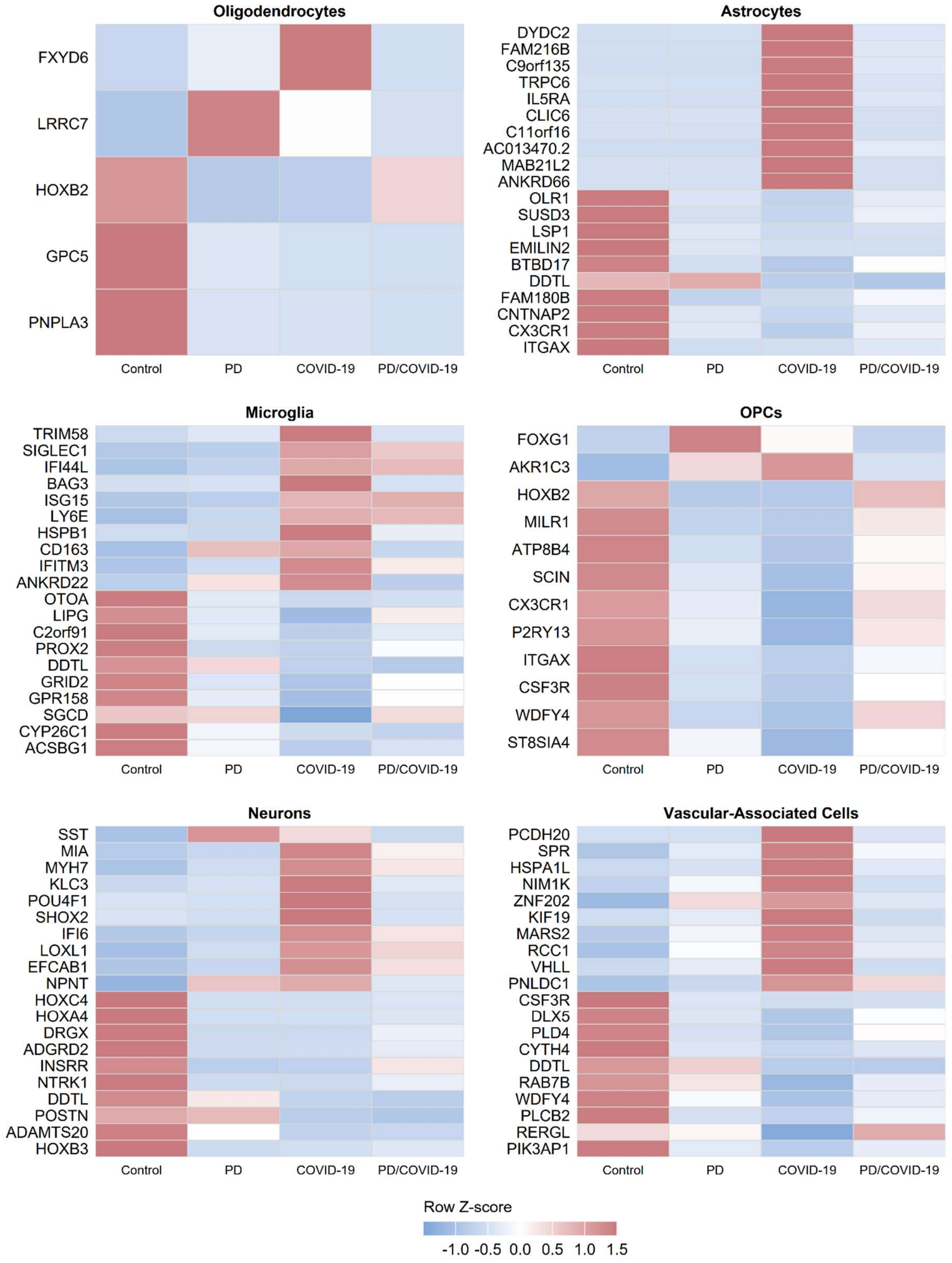


**Supplementary Fig. 3 Top differentially expressed SARS-CoV-2-associated markers.** The top differentially expressed genes ( BH-adjusted $p < 0.05$; $|\log_2FC| > 3$) identified from pairwise comparisons between the COVID-19 and Control groups are shown. Heatmap values represent row-scaled (Z-score) mean expression across all four condition groups: Control, PD , COVID-19, and PD/COVID-19, allowing cross-condition expression patterns to be assessed for genes defined by their SARS-CoV-2-associated differential expression.

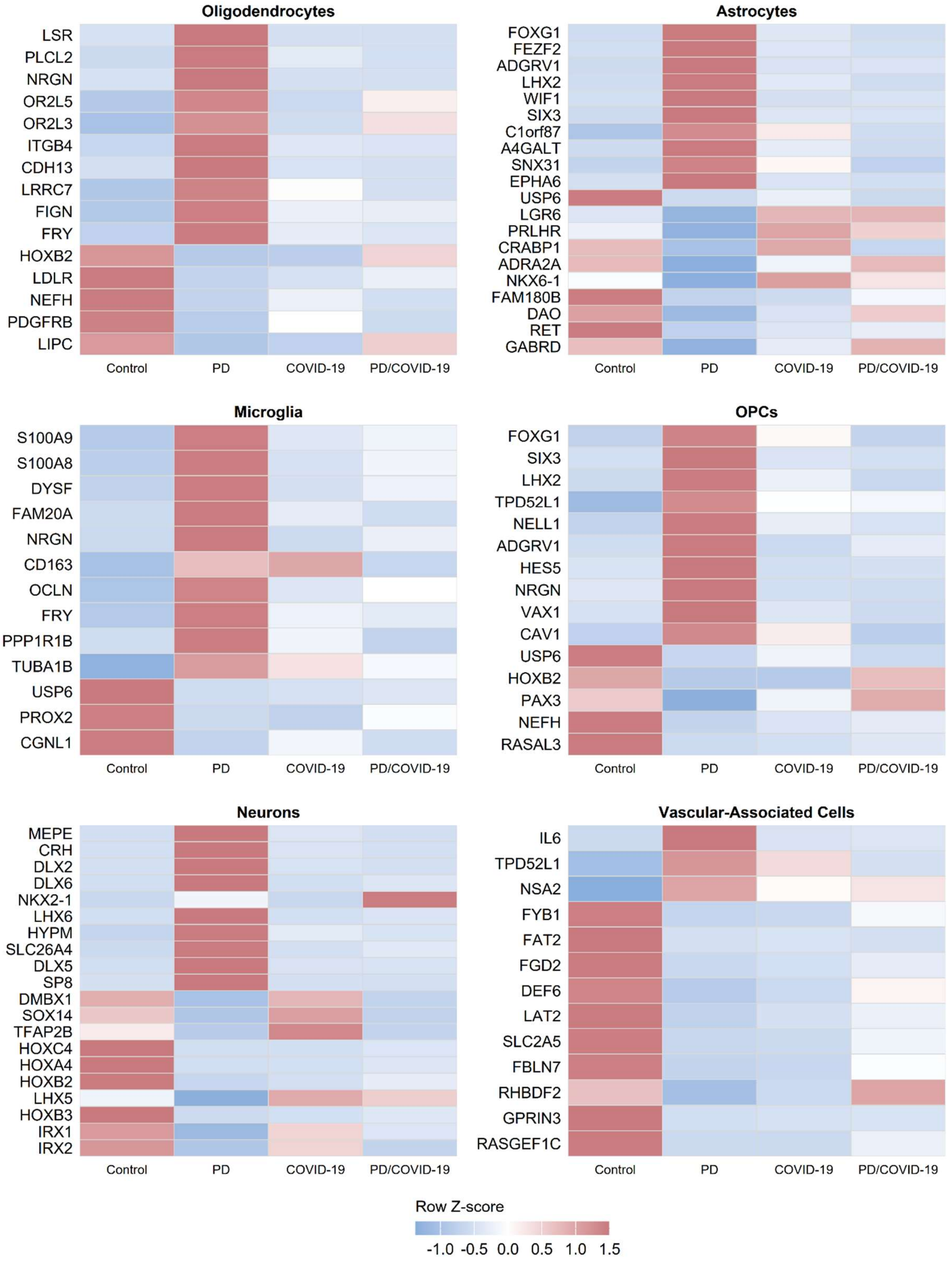


**Supplementary Fig. 4 Top differentially expressed Parkinson's Disease-associated markers.** The top differentially expressed genes (BH-adjusted $p < 0.05$; $|\log_2 FC| > 3$) identified from pairwise comparisons between the PD and Control groups. Heatmap values represent row-scaled (Z-score) mean expression across all four condition groups: Control, PD, COVID-19, and PD/COVID-19; enabling cross-condition expression patterns to be assessed for genes defined by their Parkinson's disease-associated differential expression.

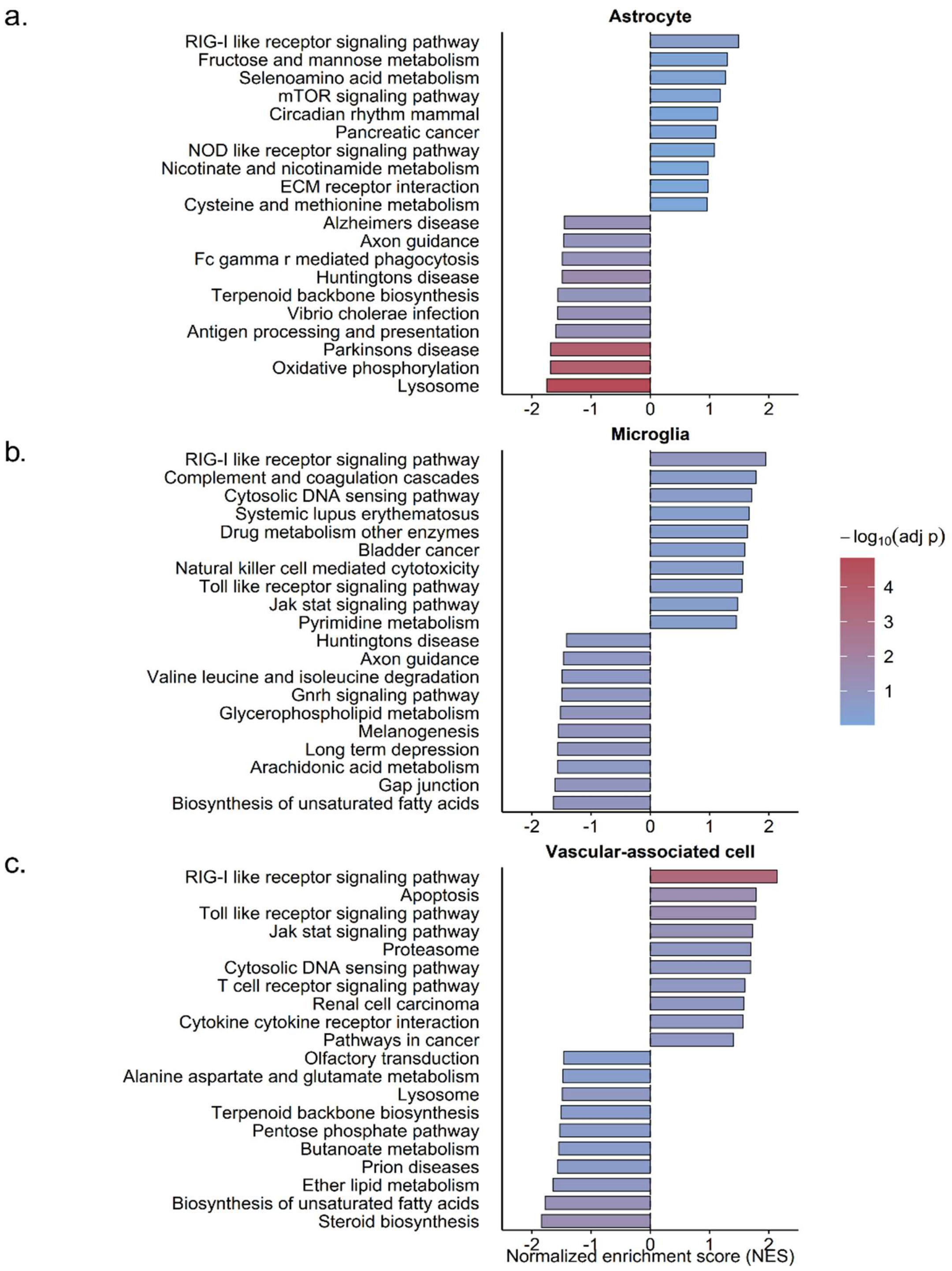


**Supplementary Fig. 5 Cell type-specific KEGG pathway enrichment in the PD and COVID-19 group.** KEGG gene set enrichment analysis was performed within astrocytes (a), microglia (b), and vascular-associated cells (c) comparing the PD/COVID-19 group relative to Control. The x-axis shows the normalized enrichment score (NES), where positive values indicate enrichment among upregulated genes and negative values indicate enrichment among downregulated genes. Dot size corresponds to pathway gene set size, and color indicates statistical significance ($-\log_{10}$ adjusted p-value)

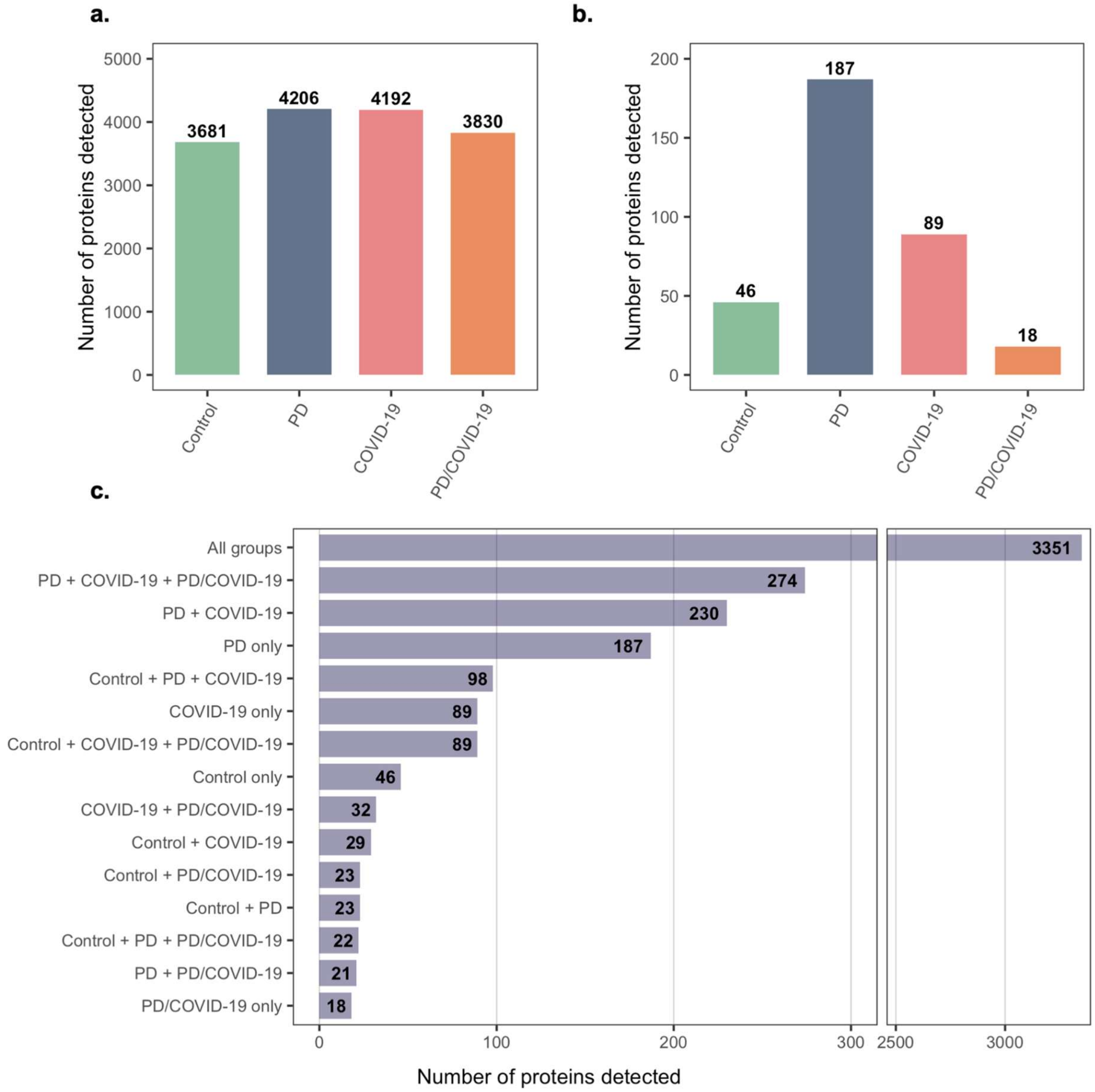


**Supplementary Figure 6. Distribution of detected proteins and shared proteomic signatures across Parkinson's disease, COVID-19, comorbid COVID-19/Parkinson's disease samples, and controls.** a. Total number of proteins detected in substantia nigra samples from Control, PD, COVID-19, and PD/COVID-19 individuals. A total of 4,532 proteins were identified across the dataset, with 3,681, 4,206, 4,192, and 3,830 proteins detected in the respective groups. b. Number of proteins uniquely detected within each experimental group. PD samples exhibited the highest number of group-specific proteins (n = 187), followed by COVID-19 (n = 89), Control (n = 46), and PD/COVID-19 (n = 18). c. Overlap analysis of detected proteins across the four groups. A large conserved core proteome comprising 3,351 proteins was shared among all groups. In addition, 274 proteins were detected in all disease-associated groups (PD, COVID-19, and PD/COVID-19) but were absent in controls, suggesting the presence of shared pathological processes associated with neurodegeneration and/or inflammatory stress responses. The x-axis break highlights overlap categories containing fewer than 300 proteins while retaining visualization of the large common proteome shared across all groups.

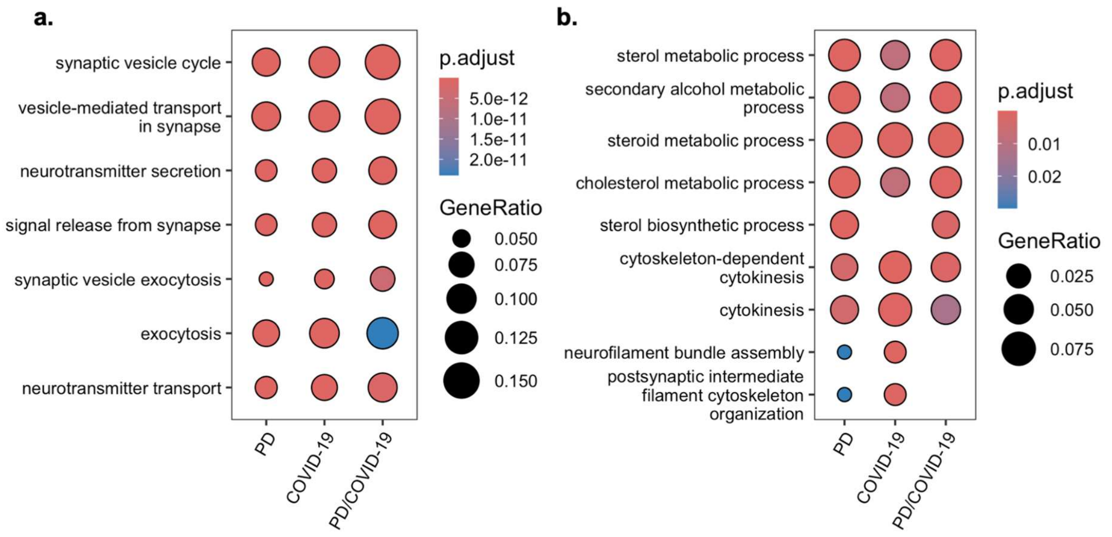


**Supplementary Figure 7. Gene ontology over-representation analysis (ORA) of differentially abundant proteins in the striatum of PD, COVID, and PD+COVID cases relative to controls.** a. Clustered comparison of significantly enriched biological process terms identified among proteins that were uniquely detected and/or significantly upregulated in the PD, COVID-19, and PD/COVID-19 groups relative to controls. Terms associated with synaptic vesicle cycling, neurotransmitter transport, exocytosis, and vesicle-mediated transport were consistently enriched across all disease groups. Numbers in parentheses indicate the number of proteins included in each enrichment analysis. Dot size represents the gene ratio, and dot colour represents the adjusted *P* value.

b. Clustered comparison of significantly enriched biological process terms identified among proteins that were uniquely detected and/or significantly downregulated in the PD, COVID-19, and PD/COVID-19 groups relative to controls. Enriched biological processes were predominantly associated with sterol, cholesterol, and steroid metabolism, together with cytoskeletal organization and cytokinesis-related pathways. Numbers in parentheses indicate the number of proteins included in each enrichment analysis. Dot size represents the gene ratio, and dot colour represents the adjusted *P* value.

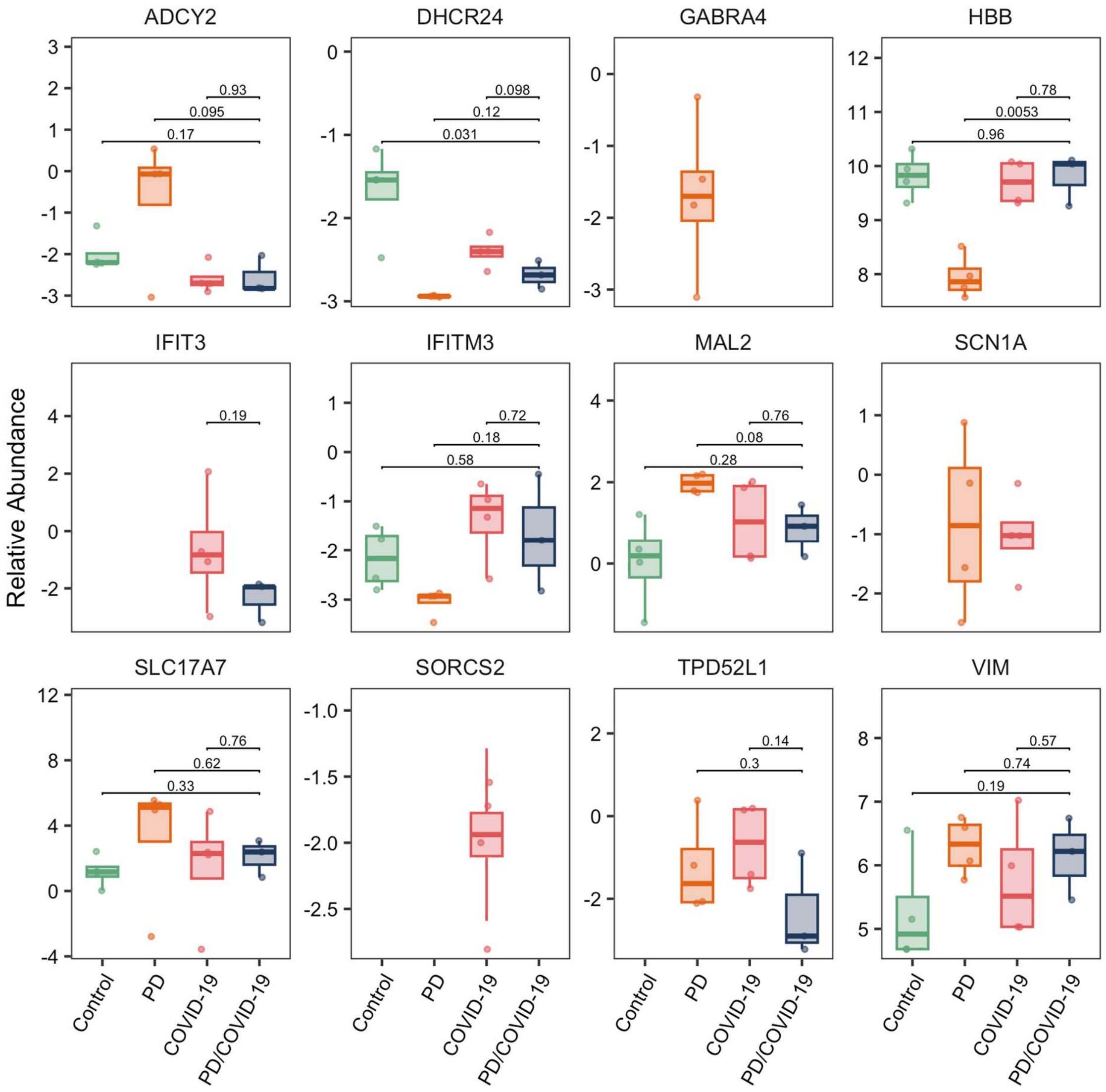


**Supplementary Figure 8. Proteomic abundance of candidates overlapping with top differentially expressed genes identified by single-nucleus RNA sequencing.** Box-and-whisker plots showing the relative abundance of proteins whose corresponding genes were identified among the top differentially expressed candidates in the single-nucleus RNA sequencing (snRNA-seq) dataset. Protein abundances are shown for Control controls, PD, COVID-19, and PD/COVID-19 groups. Boxes represent the interquartile range, center lines indicate the median, and individual points represent biological replicates. Horizontal brackets indicate pairwise group comparisons, with corresponding *P* values shown above each comparison.

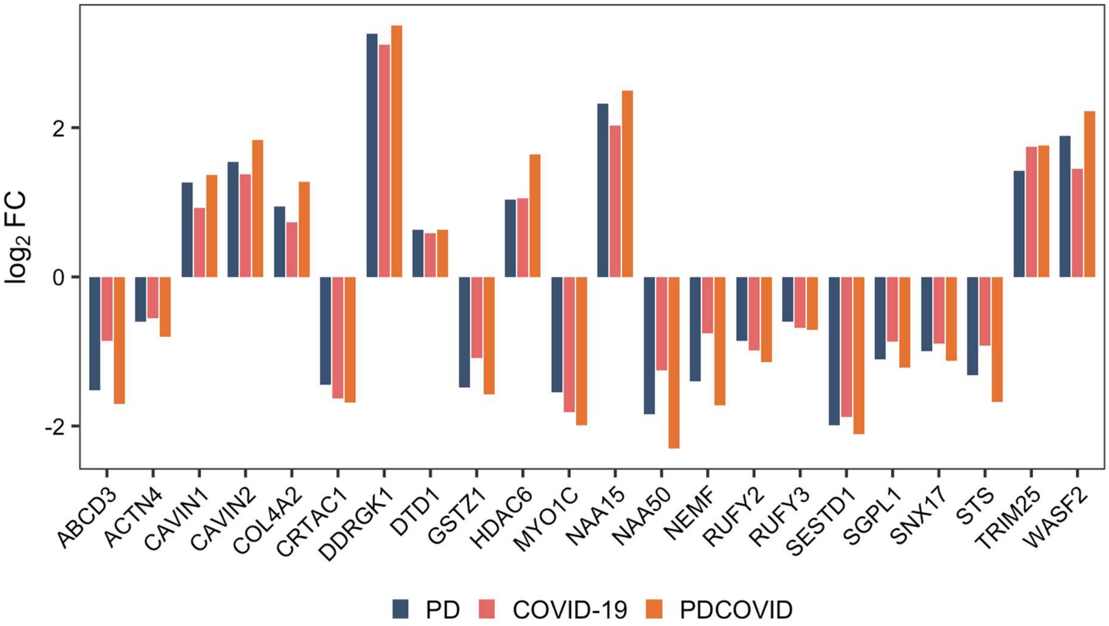


**Supplementary Figure 9. Relative abundances of proteins exhibiting amplified changes in the comorbid PD/COVID-19 condition.** Bar plots show the $\log_2$ fold changes of proteins that were differentially regulated in the same direction across the PD, COVID-19, and PD/COVID-19 groups, but displayed a greater magnitude of change in the comorbid PD/COVID-19 group than in either disease alone, consistent with an amplified comorbidity-associated effect.

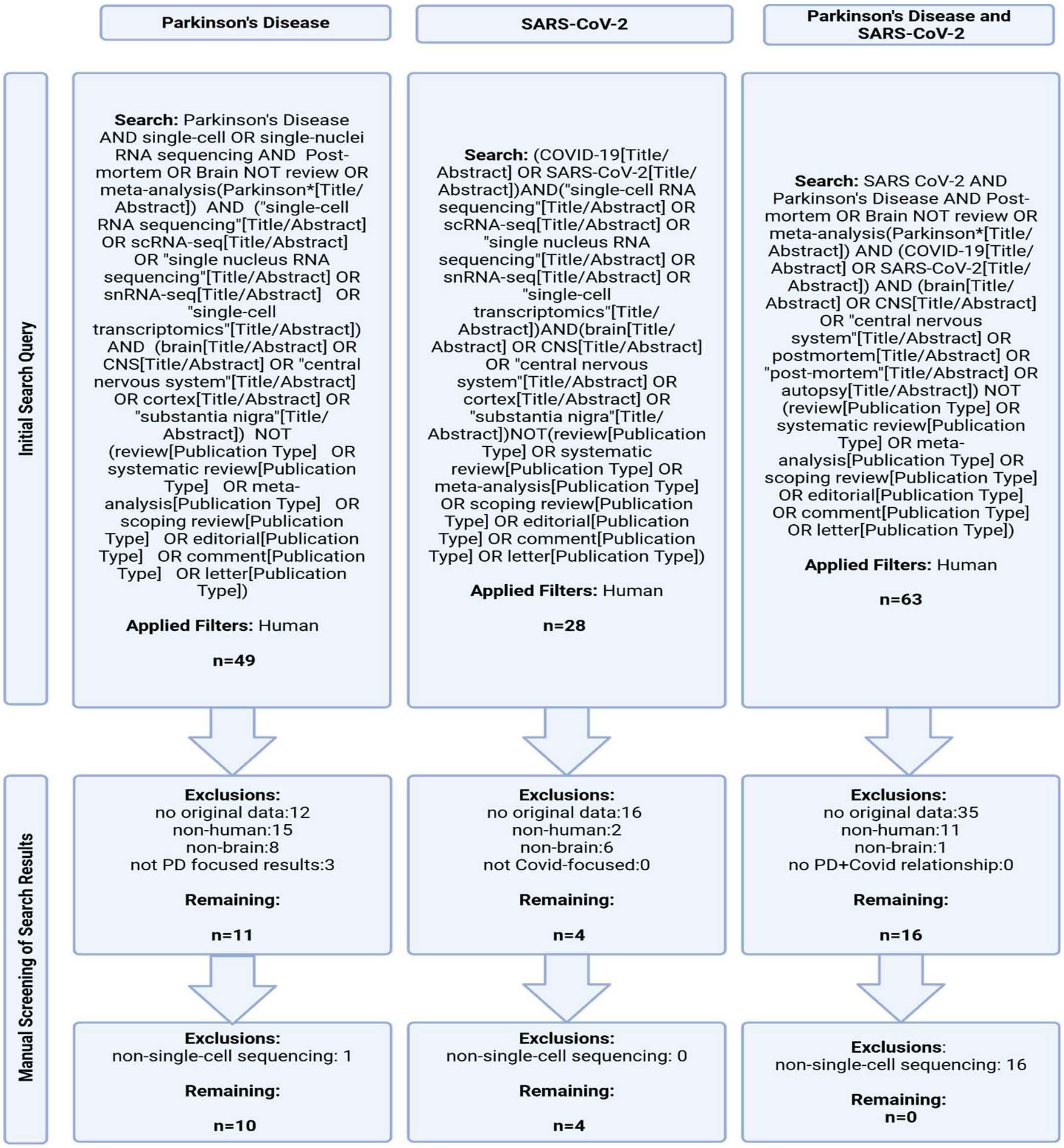


**Supplementary Fig. 10 Literature search and study selection workflow for single-nucleus RNA sequencing studies in Parkinson's disease and SARS-CoV-2.** A structured search identified primary human studies of post-mortem brain tissue in PD, SARS-CoV-2, or both. After sequential exclusion of non-human, non-brain, review, secondary, and database-derived studies, ten PD studies, four SARS-CoV-2 studies, and no combined PD/SARS-CoV-2 studies remain
